# Obstructive sleep apnea mediates genetic risk of Diabetes Mellitus: The Hispanic Community Health Study/Study of Latinos

**DOI:** 10.1101/2024.09.10.24313336

**Authors:** Yana Hrytsenko, Brian W. Spitzer, Heming Wang, Suzanne M. Bertisch, Kent Taylor, Olga Garcia-Bedoya, Alberto R Ramos, Martha L. Daviglus, Linda C Gallo, Carmen Isasi, Jianwen Cai, Qibin Qi, Carmela Alcantara, Susan Redline, Tamar Sofer

## Abstract

**Objective:** We sought to evaluate whether obstructive sleep apnea (OSA), and other sleep disorders, increase genetic risk of developing diabetes mellitus (DM).

**Research Design and Methods:** Using GWAS summary statistics from the DIAGRAM consortium and Million Veteran Program, we developed multi-ancestry Type 2 Diabetes (T2D) polygenic risk scores (T2D-PRSs) useful in admixed Hispanic/Latino individuals. We estimated the association of the T2D-PRS with cross-sectional and incident DM in the Hispanic Community Health Study/Study of Latinos (HCHS/SOL). We conducted a mediation analysis with T2D-PRSs as an exposure, incident DM as an outcome, and OSA as a mediator. Additionally, we performed Mendelian randomization (MR) analysis to assess the causal relationship between T2D and OSA.

**Results:** Of 12,342 HCHS/SOL participants, at baseline, 48.4% were normoglycemic, 36.6% were hyperglycemic, and 15% had diabetes, and 50.9% identified as female. Mean age was 41.5, and mean BMI was 29.4. T2D-PRSs was strongly associated with baseline DM and with incident DM. At baseline, a 1 SD increase in the primary T2D-PRS had DM adjusted odds ratio (OR) = 2.67, 95% CI [2.40; 2.97] and a higher incident DM rate (incident rate ratio (IRR) = 2.02, 95% CI [1.75; 2.33]). In a stratified analysis based on OSA severity categories the associations were stronger in individuals with mild OSA compared to those with moderate to severe OSA. Mediation analysis suggested that OSA mediates the T2D-PRS association with DM. In two-sample MR analysis, T2D-PRS had a causal effect on OSA, OR = 1.03, 95% CI [1.01; 1.05], and OSA had a causal effect on T2D, with OR = 2.34, 95% CI [1.59; 3.44].

**Conclusions:** OSA likely mediates genetic effects on T2D.

## Introduction

There is compelling evidence linking obstructive sleep apnea (OSA) to Diabetes Mellitus (DM), a group of metabolic diseases marked by elevated blood glucose levels due to defects in insulin secretion and utilization [1]. OSA is a common sleep-related breathing disorder characterized by repeated episodes of upper airway obstruction associated with intermittent hypoxemia and fragmented sleep-mechanisms that are implicated in impaired glucose regulation [2]. The potential pathways linking OSA and DM and evidence for a causal association have been reported previously [3–6]. In the Hispanic Community Health Study/Study of Latinos (HCHS/SOL), prospective analyses demonstrated that OSA was associated with an approximately 30% increased incidence of DM [7]. In addition to OSA, other metrics of poor sleep, including chronic partial sleep loss, have been related to DM risk [8]. Meta-analyses showed that quantity and quality of sleep, including short and long sleep durations, increase the risk of development of type 2 diabetes [9, 10]. A study in HCHS/SOL found that those with short sleep and insomnia, and long sleep without insomnia had elevated odds of diabetes prevalence [11]. DM is one of the primary risk factors for cardiovascular diseases, which in turn, is the leading cause of mortality in Hispanics/Latinos individuals [26] and thus, elucidation of the complex interplay between sleep and DM is particularly important in Hispanic/Latino populations.

Recent analysis in HCHS/SOL utilized genetic techniques to study the association of OSA with a range of phenotypes. The study showed that a polygenic risk score for OSA was associated with glycemic traits [12]; on the other hand, using GWAS summary statistics to perform a two-sample Mendelian randomization (MR) analysis, there was evidence that DM and glycemic traits were causally associated with OSA but not the other way around. These results indicate the existence of a potential bidirectional association between OSA and diabetes, which is consistent with results reported in several large cohorts [13]. However, in the manuscript that observed no causal association of OSA on diabetes [12], MR analysis was limited by use of results from a genome-wide association study of OSA that was performed in a population of Finnish Europeans and analysis of only a handful of loci obtained from analysis that did not adjust for BMI. Further, it is possible that there is a more complex association between OSA and risk of diabetes. For example, OSA may modify genetic risk of diabetes.

We hypothesized that OSA interacts with genetic risk for DM to increase DM risk. Mechanisms involved in intermittent hypoxia and fragmented sleep increase the risk of DM and are possibly related to hormonal changes caused by insufficient sleep, metabolic imbalance, chronic inflammation, or oxidative stress [14–17]. Such interactions are consistent with recent data demonstrating interactions between sleep duration and genetic loci in associations for blood pressure and lipid levels; specifically, multiple genome-wide gene-environment studies identified interactions of genomic loci with interaction with either short or long sleep duration in relation to blood pressure and lipid measures [18, 19]. It is similarly possible that OSA increases the risk of DM also via modification of genetic effects. Polygenic risk scores (PRSs) are increasingly used to summarize the genetic liability of a disease [20–29]. Given the strong association between OSA and DM, it is important to study the risk conferred by PRS for DM within the context of the “biological environment” of OSA, where we assume that individuals with OSA have potentially different tissue physiological functions compared to individuals without OSA due to hypoxia, for example.

Hispanics and Latinos are of admixed origin, with three predominant ancestral populations of European (EUR), African (AFR), and Amerindian (AMR) ancestries [30]. Thus, the genomes among groups of people of Hispanic and Latino background are a mosaic of genomic intervals each inherited from AMR, EUR, and AFR ancestries. Until recently, most studies of genetic susceptibility to DM have been performed in cohorts of European or Asian ancestry [31, 32], which may not fully represent the ancestral mosaic in Hispanic, admixed individuals. Recently, several large studies of genetic susceptibility to DM via multi-ancestry meta-analysis have identified hundreds of loci associated with type 2 diabetes [30, 31] and laid the foundation for developing PRS that are useful across diverse populations, including admixed Hispanics/Latinos. Thanks to the availability of ancestry-specific GWAS, here we develop new polygenic risk scores (PRSs) for DM that are useful for admixed individuals such as Hispanics/Latinos, and study whether their association with DM is modified by OSA or, in secondary analysis, other poor sleep phenotypes.

## Methods

We used summary statistics from published GWAS and individual-level data from the Mass General Brigham (MGB) Biobank (overview of MGB Biobank data is provided in Supplementary Note 1) to develop T2D-PRS in different ways. We used genetic data to study the associations between OSA (and, in secondary analysis, other sleep measures) and DM using data from HCHS/SOL. We applied a few approaches for analysis, including PRS, MR, and mediation analysis. Because both OSA and T2D are heavily impacted by obesity, we further considered summary statistics for PRS and MR analyses based on BMI-adjusted analyses. Table 1 summarizes the genetic data and analyses performed.

**Table 1:** Outline of the analyses performed in this study.

| PRS associations with DM at baseline |  |  |  |  |
| --- | --- | --- | --- | --- |
| Analysis | GWAS | PRS type | PRS method | Stratification categories |
| **T2D-PRS association with DM at baseline: newly developed PRSs.** | BMI-unadjusted multi-ancestry T2D GWASs: **DIAGRAM, MVP**<br>BMI-adjusted European T2D GWAS: **DIAGRAM** | mgbPRSsum<br>gapPRSsum<br>PRSsum<br>BMIadjPRS | PRS-CSx<br>PRS-CS | **Primary:** Overall dataset, OSA severity categories<br>**Secondary:** stratified by sleep phenotypes categories and by Hispanic/Latino background |
| **T2D-PRS association with DM at baseline: comparison to existing PRSs** | BMI-unadjusted multi-ancestry T2D GWASs | PGS003867_PRS<br>PGS002308_PRS (existing) | PRS-CSx<br>PRS-CS | Overall dataset |
| **OSA-PRS association with DM at baseline** | BMI-adjusted and BMI-unadjusted multi-ancestry OSA GWAS: **MVP** | Genome-wide SNPs (LDPred2) | LDPred2 | Overall dataset |
| PRS associations with incident DM |  |  |  |  |
| **T2D-PRS association with incident DM: newly developed PRSs.** | BMI-unadjusted multi-ancestry T2D GWASs: **DIAGRAM, MVP**<br>BMI-adjusted European T2D GWAS: **DIAGRAM** | mgbPRSsum<br>gapPRSsum<br>PRSsum<br>BMIadjPRS | PRS-CSx<br>PRS-CS | **Primary:** Overall dataset, OSA severity categories<br>**Secondary:** stratified by sleep phenotypes categories and by Hispanic/Latino background |
| **T2D-PRS association with incident DM: comparison to existing PRSs** | BMI-unadjusted multi-ancestry T2D GWASs | PGS003867_PRS<br>PGS002308_PRS (existing) | PRS-CSx<br>PRS-CS | Overall dataset |
| PRS associations with poor sleep phenotypes at baseline |  |  |  |  |
| **T2D-PRS association with poor sleep phenotypes** | BMI-unadjusted multi-ancestry T2D GWASs: **DIAGRAM, MVP**<br>BMI-adjusted European T2D GWAS: **DIAGRAM** | mgbPRSsum<br>gapPRSsum<br>PRSsum<br>BMIadjPRS | PRS-CSx<br>PRS-CS | OSA severity categories, sleep phenotypes categories |
| **OSA-PRS association with OSA** | BMI-unadjusted and BMI-adjusted multi-ancestry OSA GWAS: **MVP** | Genome-wide SNPs (LDPred2) | LDPred2 | OSA severity categories |
| T2D-PRS interaction analysis |  |  |  |  |
| **Interaction between T2D-PRSs and OSA severity categories** | BMI-unadjusted multi-ancestry T2D GWASs: **DIAGRAM, MVP** | mgbPRSsum<br>gapPRSsum<br>PRSsum | PRS-CSx | OSA severity categories |
| Mediation analysis |  |  |  |  |
| **Mediation analysis: T2D-PRS → OSA → incident DM** | BMI-unadjusted and BMI-adjusted multi-ancestry T2D GWASs: **DIAGRAM, MVP** | mgbPRSsum<br>gapPRSsum<br>PRSsum | PRS-CSx | OSA, REI |
| Mendelian Randomization analysis |  |  |  |  |
| Analysis | GWAS | IVs | SNPs p-value threshold | MR-method |
| **Mendelian Randomization: T2D → OSA** | BMI-unadjusted and BMI-adjusted European T2D and OSA GWASs: **DIAGRAM, MVP-White** | SNPs significantly associated with T2D | $5 \times 10^{-8}$<br>$10^{-7}$<br>$10^{-5}$ | **Primary:** IVW, MR-RAPS, MR-PRESSO<br>**Secondary:** MR-Egger, weighted median, simple mode, weighted mode, MR-RAPS. |
| **Mendelian Randomization: OSA → T2D** | BMI-unadjusted and BMI-adjusted European T2D and OSA GWASs: **DIAGRAM, MVP-White** | SNPs significantly associated with OSA | $5 \times 10^{-8}$<br>$10^{-7}$<br>$10^{-5}$ | **Primary:** IVW, MR-RAPS, MR-PRESSO<br>**Secondary:** MR-Egger, weighted median, simple mode, weighted mode, MR-RAPS. |

mgbPRSsum, gapPRSsum, and PRSsum were constructed as weighted sums of the standardized ancestry-specific PRSs for each individual. In mgbPRSsum the weights were the estimated coefficients of the three PRSs in a logistic regression of DM over the PRSs, age, sex, and 10 PCs in the MGB Biobank. In gapPRSsum the weights were individual-specific, using the individual's estimated proportions of global genetic ancestry. In PRSsum the weights were all equal to 1.
BMIadjPRS was developed based on the BMI-adjusted T2D GWAS.
PGS003867\_PRS and PGS002308\_PRS were previously developed (variants and weights obtained from the PGS catalog) and used for performance comparison from the list of effect variants and their respective effect sizes based on multi-ancestry T2D GWASs reported by [48] and [49], respectively.
OSA-PRS constructed based on previously-reported variants and weights.

### Development of polygenic risk scores for T2D

For the primary T2D-PRS developed, we used summary statistics from two large GWAS efforts of T2D: the DIAGRAM consortium [33] and the Million Veteran Program (MVP) [34], where T2D GWAS were not adjusted for BMI. Both sets of summary statistics were based on individuals from multiple populations and genetic ancestries. The DIAGRAM consortium provided summary statistics from GWAS meta-analysis in East Asian (EAS), European (EUR), and South Asian (SAS) individuals. MVP provided summary statistics from analysis of White (EUR), Black (AFR), and Hispanic (AMR) HARE (harmonized ancestry and race/ethnicity) groups. Details are provided in Supplementary Table 1. We first meta-analyzed the DIAGRAM-European and MVP-White summary statistics using inverse-variance weighted meta-analysis implemented in GWAMA [35]. Next, we used PRS-CSx [36] (global shrinkage parameter ϕ was learnt from the data, other parameters left at default) to develop ancestry-specific PRS for EUR, AFR, EAS, SAS, and AMR groups (now using population descriptors provided by the Linkage Disequilibrium reference panels implemented by PRS-CSx; specifically, we used those based on the 1000 Genomes reference panels), focusing on HapMap SNPs (as available in the PRS-CSx provided reference panel data) [37]. This resulted in a list of variants and weights for each of EUR, AFR, EAS, SAS and AMR T2D-PRSs. Because of the specific admixture patterns in HCHS/SOL individuals, we only moved forward with EUR, AFR, and AMR ancestry-specific T2D-PRSs in data analysis, as described latter.

In secondary analysis we developed T2D-PRS based on European ancestry individuals only using the DIAGRAM consortium summary statistics from a BMI-adjusted GWAS [31]. Other summary statistics from BMI-adjusted analyses are not available. Thus, we used PRS-CS to develop PRS weights from the T2D BMI-adjusted GWAS employing LD-reference panel derived from European UKBB individuals. We used the maximum Neff (total reported effective sample size) reported for this data set (157,390) as the sample size, and allowed PRS-CS to learn the global shrinkage parameter ϕ from the data. All other parameters were left at their default values. The posterior effect size estimates generated by PRS-CS are the PRS weights. This PRS is referred to as BMIadjT2D-PRS.

### The Hispanic Community Health study/Study of Latinos

The HCHS/SOL is a population-based cohort study of Hispanic/Latino adults in the United States. Individuals were recruited to the study via a multi-stage sampling design, as previously described [38, 39]. The study enrolled 16,415 adult participants (18-to 74-year-old at baseline) from four geographic areas: Bronx, NY, Chicago, IL, Miami, FL, and San Diego, CA, with enrollment between 2008-2011 (visit 1). Individuals self-identified with Hispanic/Latino backgrounds including Cuban, Central American, Dominican, Mexican, Puerto Rican, and South American. During the baseline exam (visit 1), individuals responded to various questionnaires, including sleep-related, and health measures including anthropometry, scanned medications, and fasting blood samples, were collected. HCHS/SOL participants were invited to participate in a second visit (visit 2; N = 11,623), which took place from 2014-2017, on average 6 years following visit 1.

### Sleep measures

At visit 1, participants underwent home sleep testing with an ARES Unicorder 5.2 (B-Alert, Carlsbad, CA) device within a week of their exam. The device measured nasal airflow, heart rate, snoring, body position, and oxyhemoglobin saturation. Based on the device measurements, the respiratory event index (REI) was calculated as the number of respiratory events (defined as at least 50% reduction in airflow with at least 3% desaturation for 10 seconds or more) per estimated sleep hour. OSA severity was defined based on the REI, with mild OSA defined as 15≥REI≥ 5, and moderate-to-severe OSA defined as REI≥ 15. REI<5 was considered no OSA. More information on the sleep study is provided in [40].

Other sleep phenotypes were self-reported and included insomnia, defined by the Women’s Health Initiative Insomnia Rating Scale [41] WHIIRS ≥ 10, short sleep, defined by the average sleep duration in hours ≤ 6, long sleep, defined by sleep duration > 9, excessive daytime sleepiness (EDS), defined by the Epworth Sleepiness Scale [42] ESS > 10. Sleep duration was assessed through questions regarding typical wake and bedtimes on weekdays and weekends: “What time do you usually go to bed (on weekdays/weekends)?” and “What time do you usually wake up (on weekdays/weekends)?” The average sleep duration was then calculated as a weighted average, with weekday sleep duration weighted at 5/7 and weekend sleep duration at 2/7.

### DM and incident DM outcomes

Diabetes status was ascertained based on American Diabetes Association (ADA) definition or scanned medication (at the baseline exam) or self-reported diabetes medication use (at the second exam). ADA criteria rely on laboratory tests, either fasting glucose ≥126 mg/dL, or post-OGTT glucose ≥200 mg/dL, or A1C ≥ 6.5%, or scanned/transcribed anti-diabetic medication use. Individuals had incident DM if they did not have DM at visit 1 and had DM at visit 2.

### Genotyping, imputation, and PRS construction

Consented HCHS/SOL individuals were genotyped using an Illumina custom array as previously described [30]. Quality control was performed, including checks that biological sex matched reported gender. As described in Conomos et al. 2016 [30], genetic principal components (PCs) and kinship matrix, tabulating genetic relationship between individuals, were computed using PC-AiR and PC-Relate, implemented in the GENESIS R package [43, 44]. Proportions of continental ancestry were estimated as previously reported via model-based analysis using the ADMIXTURE software [45] under the assumption of four ancestral populations (West African, European, Amerindian and East Asian). Consequently, a few individuals with East Asian ancestry were removed, and the analysis was repeated with three ancestral populations (excluding East Asian). Genome-wide imputation via the Michigan imputation server [46] was conducted using the TOPMed 2.0 imputation panel. Only variants with imputation quality R2 ≥ 0.8 and minor allele frequency ≥0.01 were used in PRS construction. All PRSs were constructed in HCHS/SOL from lists of variants, alleles, and weights, using the PRSice software [47], without any clumping and thresholding.

For primary analysis, we constructed in HCHS/SOL the three ancestry-specific PRSs developed by PRS-CSx as described earlier (EUR, AFR, and AMR). The three PRSs were standardized to have mean 0 and variance 1 in the dataset. Next, we created three PRSs for each individual, summing the three ancestry-specific PRSs in different ways: (1) gapPRSsum: a weighted sum of EUR, AFR, and AMR-specific standardized PRSs, weighted by an admixed individual’s estimated proportions of global genetic ancestry; (2) PRSsum: an unweighted sum, where the three standardized ancestry-specific PRSs were summed without any weights; and (3) mgbPRSsum: a weighted sum of the standardized ancestry-specific PRSs for each individual, where weights were computed as the estimated coefficients of the three PRSs in a logistic regression of DM over the PRSs, age, sex, and 10 genetic principal components (PCs), in the Mass General Brigham (MGB) Biobank. MGB Biobank methods are provided in Supplementary Note 1. After summing the ancestry-specific PRS, we again standardized each resulting PRS measure.

For performance comparison, we constructed two additional T2D-PRSs from the list of effect variants and their respective effect sizes based on multi-ancestry T2D GWASs reported by [48] and [49]. The list of effect variants and their effect sizes were downloaded from the PGS catalog [50]. From here on we refer to them as PGS003867_PRS and PGS002308_PRS, using the PRS identifiers from the PGS catalog. These PRSs were also standardized.

We compared each model’s prediction performance based on including only standard covariates (age, sex, BMI, study center, and genetic PCs) with models that included T2D-PRSs in addition to the above covariates. We computed the area under the receiver operating characteristic curve (AUC) and its 95% confidence interval (CI) for each model. First, we split the data into a training and testing set (90% and 10% respectively), we next fitted the models on the training set only, and then applied the R built-in function *predict* on the testing set. The AUC was measured using the *auc* function from the *Metrics* R library (version 0.1.4) on 500 “train/test” splits. We computed the mean value for the AUC and set the lower and upper bounds AUC CIs to the 2.5 and 97.5 AUC percentiles, respectively.

### Association analysis of T2D-PRS with DM and incident DM

We first verified that the T2D-PRSs were associated with DM by performing the association test between T2D-PRSs with DM at visit 1. We used survey logistic regression with DM as the outcome and adjusted for age, sex, BMI, field center and the first 5 principal components of genetic data to account for potential population stratification. This analysis used survey weights computed based on visit 1 participation. We next estimated the association of the T2D-PRSs with incident DM using analysis restricted to participants who did not have DM at visit 1, in the combined dataset and stratified by Hispanic/Latino background and by sleep phenotypes. Here we performed survey Poisson regression accounting for visit 2 survey weights and using the time between the baseline clinic visit and visit 2 as an offset. Otherwise, covariates were the same as described for the baseline DM status association analysis.

Association analyses were performed among all available individuals and restricted to strata of OSA categories. In secondary analysis, we performed association analyses between T2D-PRS and DM and incident DM, stratified by other categories defined by long and short sleep durations, insomnia, and sleepiness, and stratified (separately) by self-reported Hispanic/Latino background. To reduce the number of displayed associations, secondary analyses focused on mgbPRSsum. We chose this PRS because global ancestry proportions were available only for a genetically unrelated set of individuals, reducing power, and because unweighted sums have been performing less well than the weighted sums in previous work [51].

When it appeared that T2D-PRS association differed by sleep stratum, we also performed interaction analysis, and estimated the multiplicative interaction effect between T2D-PRSs and the relevant sleep strata.

### Association analysis of T2D-PRS with sleep phenotypes

We estimated the associations of the derived multi-ancestry T2D-PRSs with poor sleep phenotypes (OSA: primary, other phenotypes: secondary) using visit 1 data. To test the association of the T2D-PRSs with sleep phenotypes, we used survey logistic regression with baseline survey weights. We set sleep phenotype as the outcome and adjusted for age, sex, BMI, field center and the first 5 genetic PCs.

### Mediation analysis of T2D-PRS as the exposure, OSA as a mediator, and incident DM as an outcome

We performed a mediation analysis using individuals without DM at visit 1 who participated in visit 2. Here, we set mgbPRSsum as the exposure and mild-to-severe (versus no) OSA as a mediator of the mgbPRSsum effect on DM. In secondary analysis, we used also BMIadjT2D-PRS as an exposure, and moderate-to-severe OSA versus no and mild OSA, as well as the continuously measured respiratory event index (REI, log transformed to achieve approximate normality) as mediators (each separately). We used survey logistic regression (linear, for REI) to fit the mediator-exposure association model, and survey Poisson regression to fit the outcome-mediator regression (here, we used the time between visits 1 and 2 to adjust for differences in follow-up duration). All models were adjusted for age, sex, and 5 genetic PCs and used visit 2 sampling weights. We used the *mediation* R package version 4.5.0 to fit the causal mediation analysis model. Because the T2D-PRSs are continuous, we computed percent mediated effect for increasing the value of the PRS from a “control” to a “treatment” value (*control.value* and *treat.value* in the code). For a given PRS, we computed the values of quantiles (0, 0.25, 0.5, 0.75, and 1) and used these. Thus, we computed (for example) the proportion of T2D-PRS effect on increasing DM risk, mediated via increasing risk for mild-to-severe OSA, if the T2D-PRS values of all individuals in the dataset changed from a “control” value of the T2D-PRS (0.25 quantile, say) to a worse “treatment” value of the T2D-PRS (0.75 quantile, say).

### Two-sample Mendelian randomization analysis of T2D and OSA

Using summary statistics from published GWAS (not HCHS/SOL data), we performed bi-directional Mendelian randomization (MR) analysis using the *TwoSampleMR* r package version 0.5.11 to estimate the causal effect between T2D and OSA. We used BMI-adjusted and BMI-unadjusted European ancestry GWASs for both T2D and OSA [31, 52, 53]. For each exposure-outcome and BMI adjustment combinations, we selected a set of instrumental variables (IVs) from the exposure GWAS by: 1) taking the intersection of SNPs between the exposure and outcome GWASs; 2) filtering the resulting SNPs by their p-value in the exposure GWAS (p < 5×10^−8^ and, secondary, 10^−7^); 3) performing clumping of the SNPs that passed the p-value threshold using the 1000 genomes European reference panel, including only bi-allelic SNPs with MAF > 0.01, and setting clumping window as *kb* = 10000 and *r2* = 0.001. Next, we harmonized the exposure and outcome data for these SNPs to ensure that the effect of a SNP on an outcome and exposure is relative to the same allele. We performed MR analysis based on these harmonized data. The primary method was the inverse variance weighted (IVW) random effects meta-analysis. We also used MR-PRESSO [54] because it allows for evaluation of horizontal pleiotropy in multi-instrument MR (we set *NbDistribution* = 10000 and *SignifThreshold* = 0.05), and MR-RAPS because it is useful when instruments are weak (e.g. p-value<10^−7^, rather than 5×10^−8^).

## Results

### HCHS/SOL participant characteristics

Table 2 characterizes the HCHS/SOL target population, overall and stratified by OSA severity categories (no OSA, mild OSA and moderate to severe OSA). Of the HCHS/SOL target population, 50.9% were females and the mean age and BMI were 41.51 (SD = 15.05) and 29.4 (SD = 6.13) respectively. At visit 1, 48.4% of the target population was normoglycemic, 36.6% was hyperglycemic and 15% met criteria for diabetes. Focusing on the population who did not have DM (normal or hyperglycemic) at visit 1 and participated in visit 2, 8.08% of the HCHS/SOL target population had incident DM at visit 2 (Supplementary Table 2). Characteristics of the HCHS/SOL target population stratified by other sleep phenotype categories are provided in Supplementary Table 3.

**Table 2:** Characteristics of the HCHS/SOL target population at baseline overall and stratified by OSA severity categories.

| Characteristic | All | No OSA | Mild OSA | Moderate to severe OSA |
| --- | --- | --- | --- | --- |
| **N** | **12,342** | **7,563** | **2,122** | **1,270** |
| **Gender N (%)** |  |  |  |  |
| Female | 7,244 (50.9) | 4,816 (55.4) | 1,117 (42.4) | 508 (31.9) |
| Male | 5,098 (49.1) | 2,747 (44.6) | 1,005 (57.6) | 762 (68.1) |
| **Age** |  |  |  |  |
| Mean (SD) | 41.51 (15.05) | 37.87 (14.03) | 50.46 (12.83) | 52.62 (12.91) |
| **BMI** |  |  |  |  |
| Mean (SD) | 29.40 (6.13) | 28.30 (5.66) | 31.47 (6.05) | 33.71 (6.25) |
| **DM status at baseline N (%)** |  |  |  |  |
| Normoglycemic | 5,090 (48.4) | 3,742 (56.8) | 544 (28.8) | 216 (18.3) |
| Hyperglycemic | 4,839 (36.6) | 2,723 (33.1) | 996 (46.8) | 598 (47.1) |
| Diabetic | 2,413 (15.0) | 1,098 (10.1) | 582 (24.4) | 456 (34.6) |
| **DM status at visit 2 N (%)** |  |  |  |  |
| Normoglycemic | 2,462 (22.5) | 1,854 (26.9) | 258 (13.6) | 98 (7.5) |
| Hyperglycemic | 3,891 (28.6) | 2,364 (27.4) | 757 (34.8) | 392 (31.5) |
| Diabetic | 2,438 (14.7) | 1,160 (10.5) | 584 (24.0) | 445 (31.0) |
| Missing DM status/<br>did not participate in visit 2 | 3,551 (34.2) | 2,185 (35.2) | 523 (27.6) | 335 (30.0) |
| **Center N (%)** |  |  |  |  |
| Bronx | 3,221 (28.7) | 1,965 (29.0) | 530 (27.1) | 292 (25.2) |
| Chicago | 2,954 (15.1) | 1,944 (16.6) | 508 (13.9) | 305 (13.8) |
| Miami | 3,346 (31.8) | 1,833 (27.9) | 552 (34.3) | 359 (36.3) |
| San Diego | 2,821 (24.3) | 1,821 (26.5) | 532 (24.7) | 314 (24.7) |
| **Alternative healthy eating index<sup>1</sup>** |  |  |  |  |
| Mean (SD) | 47.35 (7.30) | 46.96 (7.19) | 49.49 (7.41) | 49.58 (7.55) |
| **Shift work<sup>2</sup> N (%)** |  |  |  |  |
| No | 10,023 (79.4) | 6,076 (78.2) | 1,735 (80.4) | 1,063 (83.2) |
| Yes | 2,319 (20.6) | 1,487 (21.8) | 387 (19.6) | 207 (16.8) |
| **Total physical activity<sup>3</sup>** |  |  |  |  |
| Mean (SD) | 712.47 (1079.68) | 747.51 (1077.77) | 644.78 (1002.69) | 605.72 (1106.28) |
| **Vigorous physical activity<sup>4</sup>** |  |  |  |  |
| Mean (SD) | 42.01 (95.75) | 43.54 (94.89) | 38.40 (92.85) | 35.26 (92.49) |
| **Smoking (%)** |  |  |  |  |
| Never | 7,363 (60.6) | 4,801 (64.9) | 1,175 (53.5) | 648 (52.9) |
| Former | 2,468 (17.4) | 1,284 (14.4) | 548 (25.1) | 395 (28.7) |
| Current | 2,498 (22.0) | 1,471 (20.7) | 398 (21.4) | 226 (18.4) |
| **Alcohol use** |  |  |  |  |
| Never | 2,403 (18.1) | 1,444 (17.7) | 389 (16.4) | 244 (18.9) |
| Former | 4,006 (29.7) | 2,450 (29.3) | 711 (30.0) | 428 (33.3) |
| Current | 5,929 (52.2) | 3,666 (53.0) | 1,022 (53.6) | 597 (47.8) |

<sup>1</sup>A measure of diet quality based on foods and nutrients predictive of chronic disease risk [55].
<sup>2</sup>Shift work was considered either afternoon, night, split, irregular, or rotating shift. Unemployed or dayshift is not shift work.
<sup>3</sup>The total amount of time spent doing some form of physical activity in a week (MET minutes/day) based on self-report.
<sup>4</sup>The average amount of time spent per day doing vigorous physical activity (MET minutes/day) based on self-report.

### Newly-developed T2D-PRSs are associated with DM and incident DM

Here we developed three types of multi-ancestry T2D-PRSs, including PRS constructed as unweighted sum, weighted sum with weights being ancestral proportions and weights estimated from the logistic regression based on MGB Biobank dataset (Supplementary Table 4 provides characteristics of the MGB dataset and Supplementary Note 1 provides description of the MGB Biobank dataset). Figure 1a shows the proportions of individuals by DM status and the change in their DM status over time, between visit 1 and 2 (6.03 years on average), stratified by mgbPRSsum T2D-PRS quartiles. This figure demonstrates that individuals with higher values of the PRS tend to have worse DM profiles (e.g. DM already at V1). Supplementary Figure 1 shows distribution of the three new T2D-PRSs by DM category among visits 1 and 2 participants, demonstrating that, for the three PRSs, PRS distributions are shifted across individuals grouped by DM profiles (no/normal-glycemic, pre-DM/hypergelycemic, and DM).

**Figure 1:**
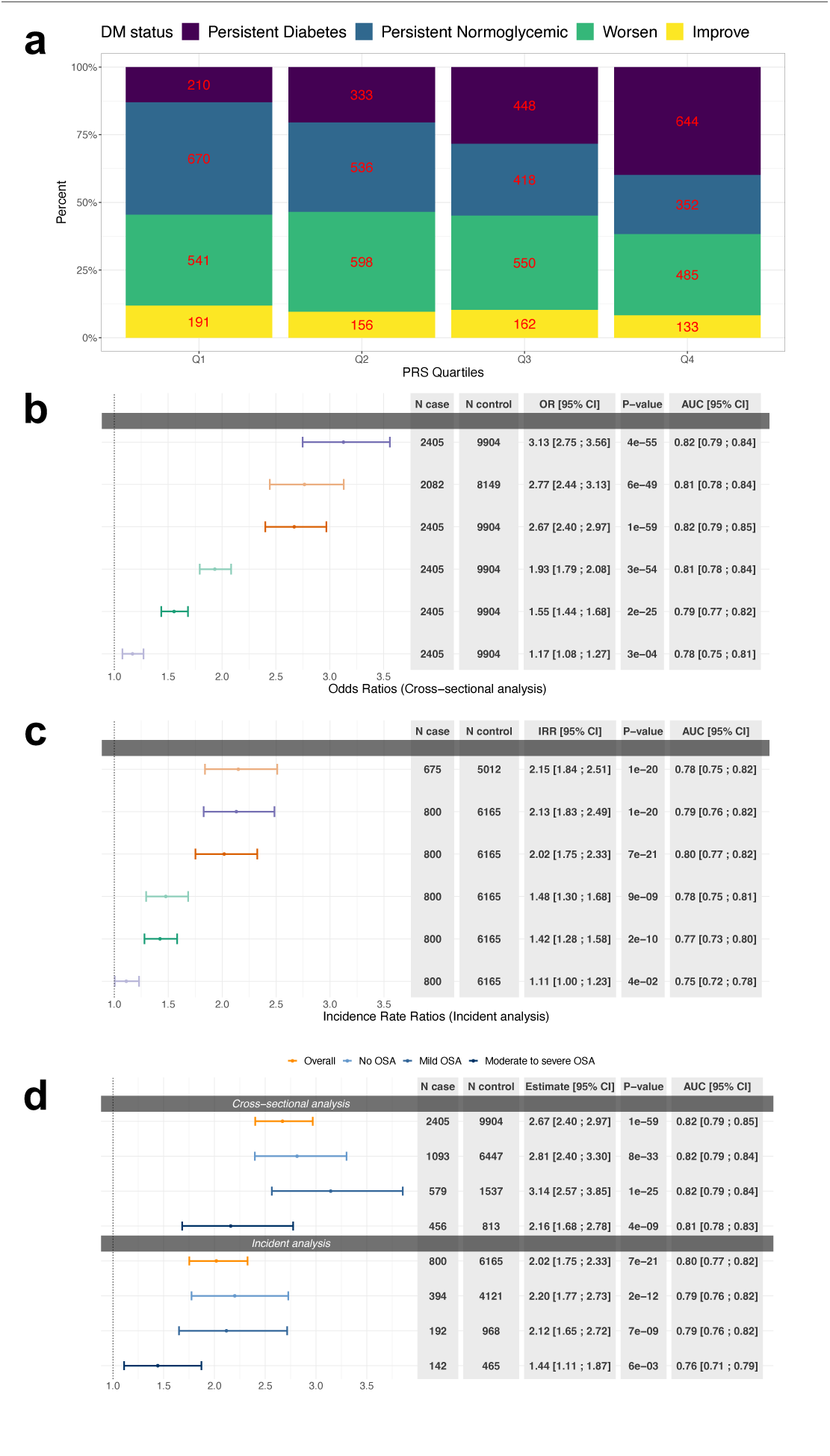
T2D-PRSs associations with DM. **Panel a)** Proportion of individuals by DM status over time (between clinic’s visit 1 and 2) stratified by T2D-PRS quartiles. Note: “Persistent” refers to having the same DM status in both visit 1 and visit 2; “Worsen” refers to change in DM status from normoglycemic at visit 1 to hyperglycemic at visit 2 or from hyperglycemic in visit 1 to diabetic at visit 2; “Improve” refers to change in DM status from hyperglycemic at visit 1 to normoglycemic at visit 2 or from diabetic in visit 1 to hyperglycemic or normoglycemic at visit 2. **b)** Estimated OR of the T2D-PRSs in association with DM at baseline in HCHS/SOL individuals with DM status at baseline **c)** Estimated IRR of the T2D-PRS in association with incident DM in individuals free of DM at baseline. **d)** Association of T2D-PRSs with DM and incident DM in HCHS/SOL individuals stratified by OSA severity levels and overall dataset. OSA severity levels were defined based on the respiratory even index (REI): mild OSA was defined as 15≥REI≥5, moderate-to-severe OSA was defined as REI≥15, and REI<5 was considered no OSA. All models were adjusted for age, sex, BMI, study center and 5 genetic PCs. OR: odds ra;os; IRR: incidence rate ra;os; AUC: Area Under the ROC (receiver opera;ng characteris;c) Curve; T2D: type 2 diabetes; PRSs: polygenic risk scores; DM: diabetes mellitus; HCHS/SOL: Hispanic Community Health Study/Study of La;nos; EDS: excessive day;me sleepiness; OSA: obstruc;ve sleep apnea.

Association analysis of T2D-PRSs with DM and incident DM in the overall dataset showed associations of all newly developed T2D-PRSs. Results across the three new T2D-DM PRSs were roughly similar. At baseline (Figure 1b), per 1 standard deviation (SD) increase of the PRS, PRSsum was associated with increased visit 1 prevalence of DM with odds ratio (OR) = 3.13, 95% confidence interval (CI) [2.75; 3.56], gapPRSsum had OR = 2.77, 95% CI [2.44; 3.13], and mgbPRSsum had OR = 2.67, 95% CI [2.4; 2.97] for mgbPRSsum. Figure 1c shows corresponding results for incident DM (N = 803) among individuals with normal glycemia and hyperglycema. Incident rate ratios (IRRs) ranged from 2.02 (PRSsum) to 2.15 (gapPRSsum) per 1 SD increase of the PRS, and all associations were highly statistically significant. In contrast, while associations with PGS002308_PRS and PGS003867_PRS (see Table 1 for description of these PRSs) were also statistically significant, effect sizes were substantially lower, with ORs of 1.55 and 1.93 for baseline DM, and IRRs of 1.42 and 1.48 for incident DM. Supplementary Figure 2 provides results from association analysis of the three new T2D-PRSs with DM and incident DM stratified by self-reported Hispanic/Latino background (characteristics of the HCHS/SOL target population stratified by background are provided in Supplementary Table 5). Here too, for each background group, the three PRSs had similar effect estimates, and all associations were statistically significant. Across groups, effect estimates from association analysis with baseline DM were highest for the South American group, with ORs of 4.44-5.63 (though wide confidence intervals given the sample size), and for incident DM the effect size estimates were highest for the Dominican group, with IRRs of 3.48-3.88.

Association analysis between the three T2D-PRSs with prevalent DM and incident DM stratified by OSA severity categories showed stronger associations in individuals with no or mild OSA compared to those with moderate-to-severe OSA, for both analyses (Figure 1d; providing results for mgbPRSsum, Supplementary Figure 3 provides results for the three T2D-PRSs). For example, in individuals with no OSA, mgbPRSsum had OR=2.81, 95% CI [2.81; 3.30] for visit 1 prevalent DM and IRR=2.2, 95% CI [1.77; 2.73] for incident DM, while in individuals with moderate-to-severe OSA it had OR=2.16, 95% CI [1.68; 2.78] and IRR=1.44 95% CI [1.11; 1.87] for visit 1 and incident DM, respectively. However, in interaction analysis models that included all individuals and interactions terms for T2D-PRSs (each in a separate model) with mild and with moderate-to-severe OSA, the interaction terms were not statistically significant, 0.21 < p-value < 0.88 (Supplementary Figure 4). We also report AUCs assessing the predictive performance of mgbPRSsum by comparing models with and without the PRS. While 95% ICs of the AUCs overlapped between models with and without the T2D-PRS, AUC values increased across all OSA severity strata and in the model using the complete population (Supplementary Figure 5). For example, the AUC in models predicting incident DM (regardless of sleep phenotype) increased from 0.75 in the model without the T2D-PRS to 0.80 in the model with the PRS.

### Evidence that OSA mediates the effect of genetic determinants of T2D on DM

The T2D-PRSs were associated with increased risk of OSA. Specifically, per 1 SD increase of mgbPRSsum, the OR for mild-to-severe OSA (versus no OSA) was 1.15, 95% CI [1.06; 1.26]. Associations with other sleep phenotypes were not statistically significant (Figure 2a). Supplementary Figure 6 shows results for the estimated association between all considered T2D-PRS (including the one constructed based on BMI-adjusted GWAS in individuals of European ancestry) and other sleep phenotypes. Results were consistent with those in Figure 2a, with the exception that the BMIadjT2D-PRS did not appear to be associated with mild-to-severe OSA (OR=1.04, p-value = 0.3).

**Figure 2:**
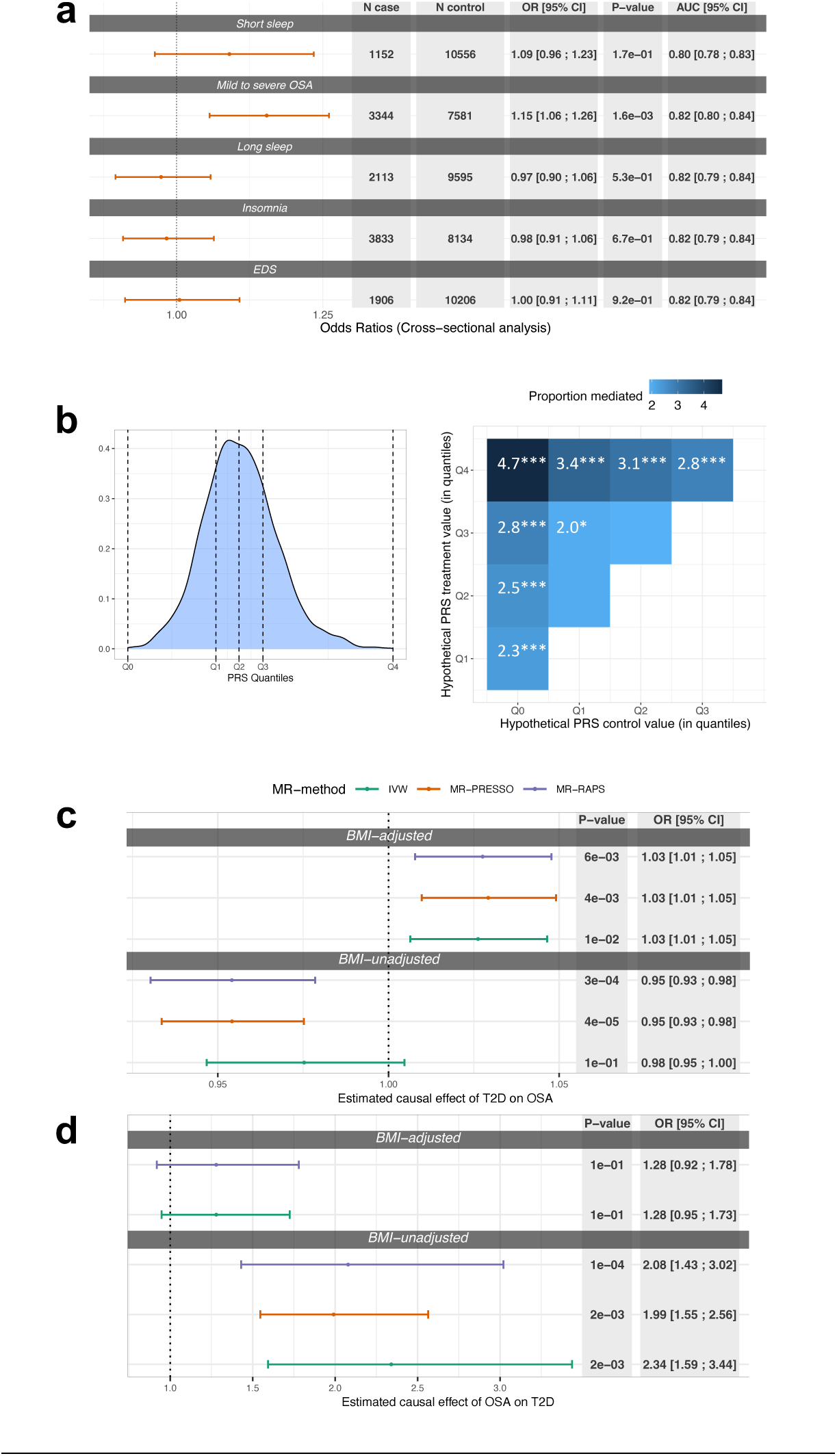
Associations of T2D-PRSs with poor sleep phenotypes, estimated mediation effect by OSA, causal effects of T2D on OSA and OSA on T2D. **Panel a)** Estimated OR of mgbPRSsum in association with poor sleep health at baseline in HCHS/SOL individuals with DM status at baseline. Results are stratified by poor sleep health categories. **b) left:** Distribution of mgbPRSsum at visit 2 (horizontal dashed lines denote quantiles of the PRS values Q_0_ – Q_4_) **right:** Estimated proportion of mediation by mild to severe OSA in the association between T2D-PRS and incident DM in individuals who participated at the second visit to a clinic (N = 6,291). Significance codes: 0 >= ‘***’ < 0.001 >= ‘**’ < 0.01 >= ‘*’ < 0.05 ‘’ < 0.1 **c)** Estimated causal effect of T2D on OSA based on SNPs selected using p-value threshold < 5*10^−8^ in BMI-adjusted and BMI-unadjusted T2D GWASs. **d)** Estimated causal effect of OSA on T2D based on SNPs selected using p-value threshold < 5*10^−8^ in BMI-adjusted and BMI-unadjusted OSA GWASs. All models were adjusted for age, sex, BMI, study center and 5 genetic PCs. T2D: type 2 diabetes; OSA: obstructive sleep apnea; IVW: inverse variance weighted; BMI: body mass index; AUC: Area Under the ROC (receiver operating characteristic) Curve; SNPs: Single-nucleotide polymorphism; GWAS: genome wide association study

In mediation analysis, OSA phenotypes mediated some of the T2D-PRS effect on DM development. Figure 2b (right panel) demonstrate the estimated percent mediated effect when using mgbPRSsum as the exposure and mild-to-severe OSA as the mediator of incident DM (outcome). In the most extreme setting, the proportion of mediation by mild-to-severe OSA versus no OSA was 4.7% when assuming that in the population the mgbPRSsum value increases from the lowest (denoted as Q0 in the figure) to the highest (denoted as Q4) observed value in the sample (see Figure 2b left panel for a visual of the Q_0_-Q_4_ PRS values cut-off points). Supplementary Figure 7 provides results from mediation analyses using mgbPRSsum as exposure and REI and moderate-to-severe OSA as mediators, showing consistent results with the primary analysis, though with lower estimated percentages of mediation. Supplementary Figure 8 provides results from mediation analyses using BMIadjT2D-PRS as exposure and the three OSA phenotypes as mediators. When using REI and mild-to-severe OSA as mediators there were statistically significant estimates of percentages of mediated risk, but not when using moderate-to-severe OSA, potentially due to lower power given smaller number of individuals with moderate-to-severe OSA.

### Genetic analysis provides evidence that T2D is a cause of OSA

Given the mediation results, we estimated the causal effect of T2D on OSA, and of OSA on T2D, using two-sample MR based on summary statistics from published GWAS of OSA and of T2D (Table 1). We identified 142 and 182 SNPs that passed the filtering steps to serve as instruments for MR of T2D on OSA based on BMI-adjusted and BMI-unadjusted T2D GWASs. Figure 2c provides results from MR of T2D on OSA, showing evidence for a weak causal association of T2D on OSA when accounting for BMI via BMI-adjusted analysis: IVW OR = 1.03, 95% CI [1.01; 1.05]; the same effect sizes were observed in sensitivity analyses using other MR methods. BMI-unadjusted anlaysis suggested no causal effect: IVW OR = 0.98, 95% CI [0.95; 1].

In MR analysis of OSA on T2D, we had 2 and 10 SNPs serving as instruments for MR based on BMI-adjusted and BMI-unadjusted primary analyses. Results are visualized in Figure 2d. BMI-unadjusted MR suggested causal effect of OSA on T2D (IVW OR=2.34, 95% CI [1.59; 3.44]) but in BMI-adjusted analysis the association weakened and was no longer statistically significant. Supplementary Figure 9 demonstrates similar results based on an analysis that utilized weak instruments (p-value<10^−7^ in the OSA GWAS). Supplementary Tables 6 and 7 provide complete lists of SNPs used in estimation of causal effect of T2D on OSA and Supplementary Tables 8 and 9 provide the same for OSA on T2D.

Because MR analysis was limited by the availability of GWAS of only European ancestry populations, and only a few strong instruments for OSA (reducing power), we also constructed OSA-PRSs in HCHS/SOL and (a) validated its association with OSA, and then (b) estimated its association with DM (noting that the interpretation of such an association is limited to prediction rather than to causality). We constructed the OSA-PRS developed using LDPred2 [56] based on the multi-ethnic GWAS of OSA in the Million Veteran Program [53], as reported in [57]. Supplementary Figure 10 demonstrates that the two OSA-PRSs (BMIadjOSA-PRS and BMIunadjOSA-PRS, based on BMI-adjusted and –unadjusted GWAS, respectively) were associated with moderate-to-severe OSA versus no-and-mild OSA, as well as with mild-to-severe OSA versus no OSA. Next, we estimated the associations of two OSA-PRSs with baseline DM. The estimated associations were similar and close to null for both BMIadjOSA and BMIunadjOSA PRSs (OR= 1.03, 1.02) and statistically insignificant (p-values >0.7). These results are provided in Supplementary Figure 11.

## Discussion

As a summary of the key clinical insights of this work: In a large Hispanic/Latino cohort using multi-ancestry T2D-PRS designed to be powerful for individuals from admixed ancestral backgrounds, the data revealed that OSA is both a causal risk factor for DM and also mediates some of the genetic risk for developing DM. This suggests that OSA is a modifiable risk factor for both existing and incident cases of DM. More specifically, we developed multi-ancestry T2D-PRSs and used them to study the relationship between OSA and DM in the HCHS/SOL. T2D-PRSs were highly associated with DM, and, contrary to our expectation, their association with DM appeared weaker in individuals with moderate-to-severe OSA compared to individuals with no or with mild OSA (though an interaction test was not statistically significant). Subsequent analysis suggested that T2D-PRS effect on DM is mediated via an increase in OSA severity.

Causal association analysis using two-sample MR suggested that T2D is causally associated with increased OSA risk when adjusting for BMI, while OSA is causally associated with increased T2D risk when not accounting for BMI. However, MR results are limited by the data availability: the strength of instruments for MR analysis and the source population of summary statistics.

We used the PRS-CSx package to develop ancestry-specific PRSs, which we then combined as sums: PRSsum, gapPRSsum, and mgbPRSsum. The three PRSs had similar performance overall (with small variations across various analyses and stratifications) and additional datasets are needed to potentially identify whether one approach is better than other approaches. Critically, gapPRSsum used individual-specific weights, depending on the estimated global ancestry proportions of an individual to sum the PRSs. The number of individuals for whom gapPRSsum (N = 10,258) was computed is lower than the sample sizes used in analysis with the other PRSs (N = 12,342), due to missing availability of estimated genetic ancestry proportions. To generalize the gapPRSsum to other populations, one needs to have both ancestry-specific PRSs and the corresponding ancestry proportion estimates for each individual in the new population. This is a limitation because these may not be available, or the specific ancestry selected (or most appropriate) for inference for a given population may not correspond to the ancestry-specific PRSs. Further, the same strategy may not be applicable for different ancestry specifications due to lack of data availability. Other PRS combination strategies may also be limited: mgbPRSsum is limited due to the use of the specific MGB population, which has different ancestry composition (and other demographic characteristics, that may impact PRS weights estimation) than other target populations. PRSsum is limited in that it places the same weight on all PRSs, while one or some may be substantially stronger than others due to the sample size in the source GWAS population. More generally, it is plausible that different PRS combination weights are suitable for different populations, just like different weights may be suitable for different individuals (as in gapPRSsum). The sample size in our study likely was too small to allow for reaching a conclusion either way, and ideally, several studies of well-defined genetic ancestry makeup would be used to assess ideal weighting strategies. In summary, the three PRSs had good performance and improved over existing PRSs, and either one can be used for future research.

Several earlier studies, including those conducted in Hispanic/Latino individuals, demonstrated evidence of the association between OSA and other measures of poor sleep health with DM [7, 8, 11, 58–61]. Here we observe a strong association between T2D-PRSs and OSA. However, there was no statistically significant evidence of the association of T2D-PRS with insomnia, short sleep, long sleep, or EDS. A meta-analysis of published studies examined the associations between OSA with pre-DM and DM, including the impact of the severity of OSA on DM. The results showed that OSA is associated with a higher risk of DM, and DM-related glycemic traits, both longitudinally [62, 63] and cross-sectionally [64]. Furthermore, results from previous studies have shown a bidirectional association between T2D and OSA [13, 15, 65, 66]. Here, we, too, observed a bidirectional association of T2D and OSA demonstrated via results of causal association in MR analysis, albeit with differences due to the effects of BMI. Our results suggest a mediation effect by OSA on the association between T2D-PRS and incident DM, which may explain the weaker T2D-PRS and DM association in the more severe OSA category. This is because an analysis restricted to individuals with mild-to-severe OSA only estimates the portion of the T2D-PRS on DM that is not mediated via mild-to-severe OSA, i.e. only the direct effect and not the total effect. While some earlier MR-based investigations of the causal effect of OSA on T2D reported no direct causal effect by OSA [67, 68], others were able to establish an association [69] suggesting the potential indirect impact of OSA on DM via BMI. These results support our findings in two-sample MR, which used published summary statistics from GWAS in individuals of European genetic ancestries, rather than the HCHS/SOL dataset. In the MR analysis causal effect of OSA on T2D was found in BMI-unadjusted analysis but not in BMI-adjusted analysis. BMI has a known causal effect on OSA [70] which may further explain our findings of the causal effect of OSA on T2D in BMI-unadjusted analysis where results may be largely influenced by BMI.

Previous publications studied the associations between sleep-related traits (including sleep duration and quality, insomnia, short and long sleep, morningness-eveningness chronotype, and others) and risk of DM. For example, [71–73] used MR analysis and established causal association between sleep and DM. However, other studies [72, 73] found no evidence of a causal association between sleep duration and the risk of DM. Also, published analyses reported significant genetic correlations of T2D with insomnia, and short and long sleep [74]. In contrast, our results, using a different analytic approach, showed weak associations between T2D-PRS and insomnia, and short and long sleep. However, only modest sample sizes and potential confounding with other health measures and health behaviors may have limited our power. For example, individuals with healthy sleep are younger individuals than others, and this may substantially affect the association results. Differences in questionnaires and measurement methods used to assess sleep phenotypes may also contribute to differences in findings between studies.

A strength of this work is that we constructed multi-ancestry T2D-PRS that factor information on an individual’s admixed ancestry and may be more suitable in analyses that are applied to samples of Hispanic/Latino individuals. Previously, it was demonstrated that PRSs are less effective in predicting outcomes for individuals whose genetic background significantly deviates from that of the participants in the original GWAS from which the scores were calculated [75, 76]. Thus, accounting for multiple genetic ancestries in the construction of multi-ancestry T2D-PRSs potentially enhances predictive power in the analysis performed on the data of Hispanic/Latino adults. Further, our study is based on the population of diverse Hispanic/Latino adults in the U.S., which is unrepresented in research. OSA was assessed via an objective, overnight sleep study, and we used a few analytic strategies, leveraging both individual-level data (PRS, mediation analyses) and summary statistics (MR) to study how DM and OSA related to each other (and other sleep phenotypes in secondary analyses). A limitation in this study is that we treated type 1 and type 2 DMs combined as DM. We were unable to distinguish between the two types of DM because the HCHS/SOL dataset only contains information on an DM status without specifying DM type. Nevertheless, given the low prevalence of T1D and the strong associations of the T2D-PRSs with both baseline DM and incident DM, which is likely mostly T2D given that our study population was, for the most part, older than age 18 years at baseline, we believe that the results are robust, in that T2D-PRSs are predictive and the relationship with OSA is well characterized.

In summary, we developed PRSs for T2D utilizing information from multiple genetic ancestries and specifically focusing on the ancestral populations of admixed Hispanic/Latino individuals in the U.S. The multi-ancestry T2D-PRSs had a strong association with baseline DM and incident DM. They were also associated with OSA, providing evidence that OSA mediates some of the T2D risk conferred by underlying genetic factors. This work extends our current knowledge of understanding the effect of OSA on the risk of developing DM later in life, given an individual’s genetic predisposition to T2D.

## Data availability statement

Ancestry-specific summary statistics from GWAS of DM published in 2022 (Mahajan et al) in the DIAGRAM consortium were downloaded from https://diagram-consortium.org/downloads.html. HARE-group specific summary statistics from GWAS of DM in MVP published in 2020 (Vujkovic et al) were downloaded by dbGaP application to study accession phs001672. SNPs and weights for ancestry-specific T2D-PRSs are provided in the GitHub repository https://github.com/YanaHrytsenko/DM_PRS_OSA_mediation.

## Code availability statement

Code used for analysis in this paper is publicly available on the GitHub repository https://github.com/YanaHrytsenko/DM_PRS_OSA_mediation.

## Ethics statement

The HCHS/SOL was approved by the institutional review boards (IRBs) at each field center, where all participants gave written informed consent, and by the Non-Biomedical IRB at the University of North Carolina at Chapel Hill, to the HCHS/SOL Data Coordinating Center. All IRBs approving the HCHS/SOL study are: Non-Biomedical IRB at the University of North Carolina at Chapel Hill. Chapel Hill, NC; Einstein IRB at the Albert Einstein College of Medicine of Yeshiva University. Bronx, NY; IRB at Office for the Protection of Research Subjects (OPRS), University of Illinois at Chicago. Chicago, IL; Human Subject Research Office, University of Miami. Miami, FL; Institutional Review Board of San Diego State University, San Diego, CA. All methods and analyses of HCHS/ SOL participants’ materials and data were carried out in accordance with human subject research guidelines and regulations. This work was approved by the Mass General Brigham IRB and by the Beth Israel Deaconess Medical Center Committee on Clinical Investigations.

## Acknowledgements

The authors thank the staff and participants of HCHS/SOL for their important contributions. Investigators website – http://www.cscc.unc.edu/hchs/.

The work was supported by National Heart Lung and Blood Institute (NHLBI) grants R01HL161012 to T.S. and R35HL135818 to S.R., and National Institute on Aging grant R01AG080598 to T.S. The Hispanic Community Health Study/Study of Latinos is a collaborative study supported by contracts from the National Heart, Lung, and Blood Institute (NHLBI) to the University of North Carolina (HHSN268201300001I / N01-HC-65233), University of Miami (HHSN268201300004I / N01-HC-65234), Albert Einstein College of Medicine (HHSN268201300002I / N01-HC-65235), University of Illinois at Chicago (HHSN268201300003I / N01-HC-65236 Northwestern Univ), and San Diego State University (HHSN268201300005I / N01-HC-65237). The following Institutes/Centers/Offices have contributed to the HCHS/SOL through a transfer of funds to the NHLBI: National Institute on Minority Health and Health Disparities, National Institute on Deafness and Other Communication Disorders, National Institute of Dental and Craniofacial Research, National Institute of Diabetes and Digestive and Kidney Diseases, National Institute of Neurological Disorders and Stroke, NIH Institution-Office of Dietary Supplements.

## References

1. American Diabetes, A., Diagnosis and classification of diabetes mellitus. Diabetes Care, 2004. 27 **Suppl 1**: p. S5–S10.

2. Senaratna, C.V., et al., Prevalence of obstructive sleep apnea in the general population: A systematic review. Sleep Med Rev, 2017. 34: p. 70–81.

3. Kono, M., et al., Obstructive sleep apnea syndrome is associated with some components of metabolic syndrome. Chest, 2007. 131(5): p. 1387–92.

4. Foster, G.D., et al., Obstructive sleep apnea among obese patients with type 2 diabetes. Diabetes Care, 2009. 32(6): p. 1017–9.

5. Pamidi, S., R.S. Aronsohn, and E. Tasali, Obstructive sleep apnea: role in the risk and severity of diabetes. Best Pract Res Clin Endocrinol Metab, 2010. 24(5): p. 703–15.

6. Nagayoshi, M., et al., Obstructive sleep apnea and incident type 2 diabetes. Sleep Med, 2016. 25: p. 156–161.

7. Li, X., et al., Associations of Sleep-disordered Breathing and Insomnia with Incident Hypertension and Diabetes. The Hispanic Community Health Study/Study of Latinos. Am J Respir Crit Care Med, 2021. 203(3): p. 356–365.

8. Knutson, K.L. and E. Van Cauter, Associations between sleep loss and increased risk of obesity and diabetes. Ann N Y Acad Sci, 2008. 1129: p. 287–304.

9. Cappuccio, F.P., et al., Quantity and quality of sleep and incidence of type 2 diabetes: a systematic review and meta-analysis. Diabetes Care, 2010. 33(2): p. 414–20.

10. Shan, Z., et al., Sleep duration and risk of type 2 diabetes: a meta-analysis of prospective studies. Diabetes Care, 2015. 38(3): p. 529–37.

11. Cespedes, E.M., et al., Joint associations of insomnia and sleep duration with prevalent diabetes: The Hispanic Community Health Study/Study of Latinos (HCHS/SOL). J Diabetes, 2016. 8(3): p. 387–97.

12. Zhang, Y., et al., Genetic determinants of cardiometabolic and pulmonary phenotypes and obstructive sleep apnoea in HCHS/SOL. EBioMedicine, 2022. 84: p. 104288.

13. Huang, T., et al., A Population-Based Study of the Bidirectional Association Between Obstructive Sleep Apnea and Type 2 Diabetes in Three Prospective U.S. Cohorts. Diabetes Care, 2018. 41(10): p. 2111–2119.

14. Briancon-Marjollet, A., et al., The impact of sleep disorders on glucose metabolism: endocrine and molecular mechanisms. Diabetol Metab Syndr, 2015. 7: p. 25.

15. Chattu, V.K., et al., The Interlinked Rising Epidemic of Insufficient Sleep and Diabetes Mellitus. Healthcare (Basel), 2019. 7(1).

16. Song, S.O., et al., Metabolic Consequences of Obstructive Sleep Apnea Especially Pertaining to Diabetes Mellitus and Insulin Sensitivity. Diabetes Metab J, 2019. 43(2): p. 144–155.

17. Pallayova, M., D. Banerjee, and S. Taheri, Novel insights into metabolic sequelae of obstructive sleep apnoea: a link between hypoxic stress and chronic diabetes complications. Diabetes Res Clin Pract, 2014. 104(2): p. 197–205.

18. Noordam, R., et al., Multi-ancestry sleep-by-SNP interaction analysis in 126,926 individuals reveals lipid loci stratified by sleep duration. Nat Commun, 2019. 10(1): p. 5121.

19. Wang, H., et al., Multi-ancestry genome-wide gene-sleep interactions identify novel loci for blood pressure. Mol Psychiatry, 2021. 26(11): p. 6293–6304.

20. Lewis, C.M. and E. Vassos, Polygenic risk scores: from research tools to clinical instruments. Genome Med, 2020. 12(1): p. 44.

21. Kurniansyah, N., et al., A multi-ethnic polygenic risk score is associated with hypertension prevalence and progression throughout adulthood. Nat Commun, 2022. 13(1): p. 3549.

22. Kurniansyah, N., et al., Evaluating the use of blood pressure polygenic risk scores across race/ethnic background groups. Nat Commun, 2023. 14(1): p. 3202.

23. Elgart, M., et al., Non-linear machine learning models incorporating SNPs and PRS improve polygenic prediction in diverse human populations. Commun Biol, 2022. 5(1): p. 856.

24. Krapohl, E., et al., Multi-polygenic score approach to trait prediction. Mol Psychiatry, 2018. 23(5): p. 1368–1374.

25. Sinnott-Armstrong, N., et al., Genetics of 35 blood and urine biomarkers in the UK Biobank. Nat Genet, 2021. 53(2): p. 185–194.

26. Pain, O., et al., Evaluation of polygenic prediction methodology within a reference-standardized framework. PLOS Genetics, 2021. 17(5): p. e1009021.

27. Abraham, G., et al., Genomic risk score offers predictive performance comparable to clinical risk factors for ischaemic stroke. Nat Commun, 2019. 10(1): p. 5819.

28. Rodriguez, V., et al., Use of multiple polygenic risk scores for distinguishing schizophrenia-spectrum disorder and affective psychosis categories in a first-episode sample; the EU-GEI study. Psychol Med, 2023. 53(8): p. 3396–3405.

29. Meisner, A., et al., Combined Utility of 25 Disease and Risk Factor Polygenic Risk Scores for Stratifying Risk of All-Cause Mortality. Am J Hum Genet, 2020. 107(3): p. 418–431.

30. Conomos, M.P., et al., Genetic Diversity and Association Studies in US Hispanic/Latino Populations: Applications in the Hispanic Community Health Study/Study of Latinos. Am J Hum Genet, 2016. 98(1): p. 165–84.

31. Mahajan, A., et al., Fine-mapping type 2 diabetes loci to single-variant resolution using high-density imputation and islet-specific epigenome maps. Nat Genet, 2018. 50(11): p. 1505–1513.

32. Suzuki, K., et al., Identification of 28 new susceptibility loci for type 2 diabetes in the Japanese population. Nat Genet, 2019. 51(3): p. 379–386.

33. Mahajan, A., et al., Multi-ancestry genetic study of type 2 diabetes highlights the power of diverse populations for discovery and translation. Nat Genet, 2022. 54(5): p. 560–572.

34. Vujkovic, M., et al., Discovery of 318 new risk loci for type 2 diabetes and related vascular outcomes among 1.4 million participants in a multi-ancestry meta-analysis. Nat Genet, 2020. 52(7): p. 680–691.

35. Magi, R. and A.P. Morris, GWAMA: software for genome-wide association meta-analysis. BMC Bioinformatics, 2010. 11: p. 288.

36. Ruan, Y., et al., Improving polygenic prediction in ancestrally diverse populations. Nat Genet, 2022. 54(5): p. 573–580.

37. International HapMap, C., The International HapMap Project. Nature, 2003. 426(6968): p. 789–96.

38. Sorlie, P.D., et al., Design and implementation of the Hispanic Community Health Study/Study of Latinos. Ann Epidemiol, 2010. 20(8): p. 629–41.

39. Lavange, L.M., et al., Sample design and cohort selection in the Hispanic Community Health Study/Study of Latinos. Ann Epidemiol, 2010. 20(8): p. 642–9.

40. Redline, S., et al., Sleep-disordered breathing in Hispanic/Latino individuals of diverse backgrounds. The Hispanic Community Health Study/Study of Latinos. Am J Respir Crit Care Med, 2014. 189(3): p. 335–44.

41. Levine, D.W., et al., Validation of the Women’s Health Initiative Insomnia Rating Scale in a multicenter controlled clinical trial. Psychosom Med, 2005. 67(1): p. 98–104.

42. Johns, M.W., A new method for measuring daytime sleepiness: the Epworth sleepiness scale. Sleep, 1991. 14(6): p. 540–5.

43. Conomos, M.P., et al., Model-free Estimation of Recent Genetic Relatedness. Am J Hum Genet, 2016. 98(1): p. 127–48.

44. Gogarten, S.M., et al., Genetic association testing using the GENESIS R/Bioconductor package. Bioinformatics, 2019.

45. Alexander, D.H., J. Novembre, and K. Lange, Fast model-based estimation of ancestry in unrelated individuals. Genome Res, 2009. 19(9): p. 1655–64.

46. Das, S., et al., Next-generation genotype imputation service and methods. Nat Genet, 2016. 48(10): p. 1284–1287.

47. Euesden, J., C.M. Lewis, and P.F. O’Reilly, PRSice: Polygenic Risk Score software. Bioinformatics, 2015. 31(9): p. 1466–8.

48. Shim, I., et al., Clinical utility of polygenic scores for cardiometabolic disease in Arabs. Nat Commun, 2023. 14(1): p. 6535.

49. Ge, T., et al., Development and validation of a trans-ancestry polygenic risk score for type 2 diabetes in diverse populations. Genome Med, 2022. 14(1): p. 70.

50. Lambert, S.A., et al., The Polygenic Score Catalog as an open database for reproducibility and systematic evaluation. Nat Genet, 2021. 53(4): p. 420–425.

51. Kurniansyah, N., et al., Evaluating the use of blood pressure polygenic risk scores across race/ethnic background groups. Nat Commun, 2023. 14(1): p. 3202.

52. Mahajan, A., et al., Multi-ancestry genetic study of type 2 diabetes highlights the power of diverse populations for discovery and translation. Nat Genet, 2022. 54(5): p. 560–572.

53. Sofer, T., et al., Genome-wide association study of obstructive sleep apnoea in the Million Veteran Program uncovers genetic heterogeneity by sex. EBioMedicine, 2023. 90: p. 104536.

54. Verbanck, M., et al., Detection of widespread horizontal pleiotropy in causal relationships inferred from Mendelian randomization between complex traits and diseases. Nat Genet, 2018. 50(5): p. 693–698.

55. Chiuve, S.E., et al., Alternative dietary indices both strongly predict risk of chronic disease. J Nutr, 2012. 142(6): p. 1009–18.

56. Privé, F., J. Arbel, and B.J. Vilhjálmsson, LDpred2: better, faster, stronger. Bioinformatics, 2021. 36(22-23): p. 5424–5431.

57. Huang, Y.-J., et al., The expected polygenic risk score (ePRS) framework: an equitable metric for quantifying polygenetic risk via modeling of ancestral makeup. medRxiv, 2024: p. 2024.03.05.24303738.

58. Khalil, M., et al., The association between sleep and diabetes outcomes – A systematic review. Diabetes Res Clin Pract, 2020. 161: p. 108035.

59. Ogilvie, R.P. and S.R. Patel, The Epidemiology of Sleep and Diabetes. Curr Diab Rep, 2018. 18(10): p. 82.

60. Antza, C., et al., The links between sleep duration, obesity and type 2 diabetes mellitus. J Endocrinol, 2021. 252(2): p. 125–141.

61. Li, Z.H., et al., Association of sleep and circadian patterns and genetic risk with incident type 2 diabetes: a large prospective population-based cohort study. Eur J Endocrinol, 2021. 185(5): p. 765–774.

62. Reichmuth, K.J., et al., Association of sleep apnea and type II diabetes: a population-based study. Am J Respir Crit Care Med, 2005. 172(12): p. 1590–5.

63. Muraki, I., et al., Nocturnal intermittent hypoxia and the development of type 2 diabetes: the Circulatory Risk in Communities Study (CIRCS). Diabetologia, 2010. 53(3): p. 481–8.

64. Wang, C., et al., Obstructive sleep apnea, prediabetes and progression of type 2 diabetes: A systematic review and meta-analysis. J Diabetes Investig, 2022. 13(8): p. 1396–1411.

65. Subramanian, A., et al., Risk of Incident Obstructive Sleep Apnea Among Patients With Type 2 Diabetes. Diabetes Care, 2019. 42(5): p. 954–963.

66. Gleeson, M. and W.T. McNicholas, Bidirectional relationships of comorbidity with obstructive sleep apnoea. Eur Respir Rev, 2022. 31(164).

67. Ding, X., et al., Mendelian randomization reveals no associations of genetically-predicted obstructive sleep apnea with the risk of type 2 diabetes, nonalcoholic fatty liver disease, and coronary heart disease. Front Psychiatry, 2023. 14: p. 1068756.

68. Wang, J., et al., Causal associations of sleep apnea and snoring with type 2 diabetes and glycemic traits and the role of BMI. Obesity (Silver Spring), 2023. 31(3): p. 652–664.

69. Shen, Y., et al., How is Obstructive Sleep Apnea Associated with High Blood Pressure and Diabetes Mellitus Type 2? Clues from a Two-Step Mendelian Randomized Study. Nat Sci Sleep, 2023. 15: p. 749–765.

70. Strausz, S., et al., Genetic analysis of obstructive sleep apnoea discovers a strong association with cardiometabolic health. Eur Respir J, 2021. 57(5).

71. Xiuyun, W., et al., Network Mendelian randomization study: exploring the causal pathway from insomnia to type 2 diabetes. BMJ Open Diabetes Res Care, 2022. 10(1).

72. Ma, Y., et al., Integrative Identification of Genetic Loci Jointly Influencing Diabetes-Related Traits and Sleep Traits of Insomnia, Sleep Duration, and Chronotypes. Biomedicines, 2022. 10(2).

73. Gao, X., et al., Investigating Causal Relations Between Sleep-Related Traits and Risk of Type 2 Diabetes Mellitus: A Mendelian Randomization Study. Front Genet, 2020. 11: p. 607865.

74. Lasconi, C., et al., Variant-to-gene-mapping analyses reveal a role for pancreatic islet cells in conferring genetic susceptibility to sleep-related traits. Sleep, 2022. 45(8).

75. Martin, A.R., et al., Clinical use of current polygenic risk scores may exacerbate health disparities. Nat Genet, 2019. 51(4): p. 584–591.

76. Lee, J.J., et al., Gene discovery and polygenic prediction from a genome-wide association study of educational attainment in 1.1 million individuals. Nat Genet, 2018. 50(8): p. 1112–1121.

