## Supplementary Material for "Obstructive sleep apnea mediates genetic risk of Diabetes Mellitus: The Hispanic Community Health Study/Study of Latinos"

#### Table of Contents

***Supplementary Figures*** **2**

**Supplementary Figure 1: Distribution of multi-ancestry T2D-PRSs by DM category between visit 1 and 2 to the clinic.** **2**

**Supplementary Figure 2: Association of multi-ancestry T2D-PRSs with DM and incident DM stratified by self-reported ancestral background.** **3**

**Supplementary Figure 3: T2D-PRSs associations with DM stratified by OSA categories and overall dataset.** **4**

**Supplementary Figure 4: Interaction between multi-ancestry T2D-PRSs and OSA phenotypes in association with incident DM.** **5**

**Supplementary Figure 5: Comparison of model performance for T2D-PRSs association with DM and incident DM using covariates and covariates plus T2D-PRS model.** **5**

**Supplementary Figure 6: Association of multi-ancestry T2D-PRSs with poor sleep health.** **6**

**Supplementary Figure 7: Mediation effect of OSA on associations of T2D-PRS with DM (BMI-unadjusted analysis).** **7**

**Supplementary Figure 8: Mediation effect of OSA on associations of T2D-PRS with DM (BMI-adjusted analysis).** **8**

**Supplementary Figure 9: Estimated causal effects of OSA on T2D.** **9**

**Supplementary Figure 10: Association of OSA-PRS with OSA at baseline** **10**

**Supplementary Figure 11: Association of OSA-PRS with DM at baseline** **10**

***Supplementary Note 1: the Mass General Brigham Biobank*** ***11***

***Supplementary Tables*** ***16***

**Supplementary Table 1: GWAS summary statistics used for T2D-PRS development.** **16**

**Supplementary Table 2: Characteristics of HCHS/SOL target population with no DM at baseline stratified by sleep phenotype categories.** **17**

**Supplementary Table 3: Characteristics of HCHS/SOL target population at baseline stratified by sleep phenotype categories.** **18**

**Supplementary Table 4: Characteristics of MGB dataset stratified by T2D status.** **19**

**Supplementary Table 5: Characteristics of HCHS/SOL target population stratified by self-reported Hispanic background.** **20**

***Supplementary References*** ***21***

### Supplementary Figures

#### Supplementary Figure 1: Distribution of multi-ancestry T2D-PRSs by DM category between visit 1 and 2 to the clinic.

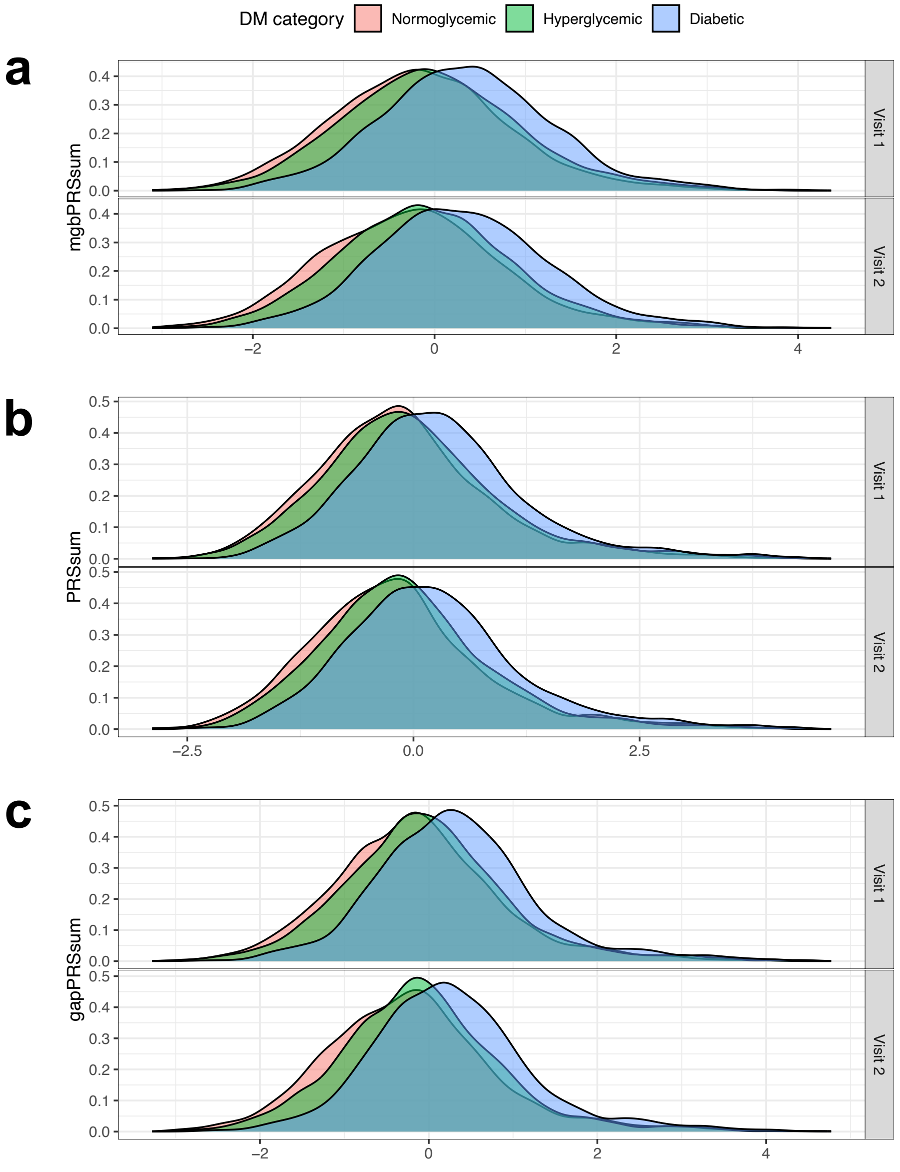


Distribution of multi-ancestry T2D-PRSs in HCHS/SOL individuals by DM category between visit 1 and 2 to the clinic. Panel a) distribution of the mgbPRSsum (visit 1: Normoglycemic N = 5,090, Hyperglycemic N = 4,839, Diabetic N = 2,413; visit 2: Normoglycemic N = 2,462, Hyperglycemic N = 3,891, Diabetic N = 2,438); b) distribution of the PRSsum (visit 1: Normoglycemic N = 5,090, Hyperglycemic N = 4,839, Diabetic N = 2,413; visit 2: Normoglycemic N = 2,462, Hyperglycemic N = 3,891, Diabetic N = 2,438); c) distribution of the gapPRSsum (visit 1: Normoglycemic N = 4,058, Hyperglycemic N = 4,111, Diabetic N = 2,089; visit 2: Normoglycemic N = 1,922, Hyperglycemic N = 3,257, Diabetic N = 2,077);

Supplementary Figure 2: Association of multi-ancestry T2D-PRSs with DM and incident DM stratified by self-reported ancestral background.

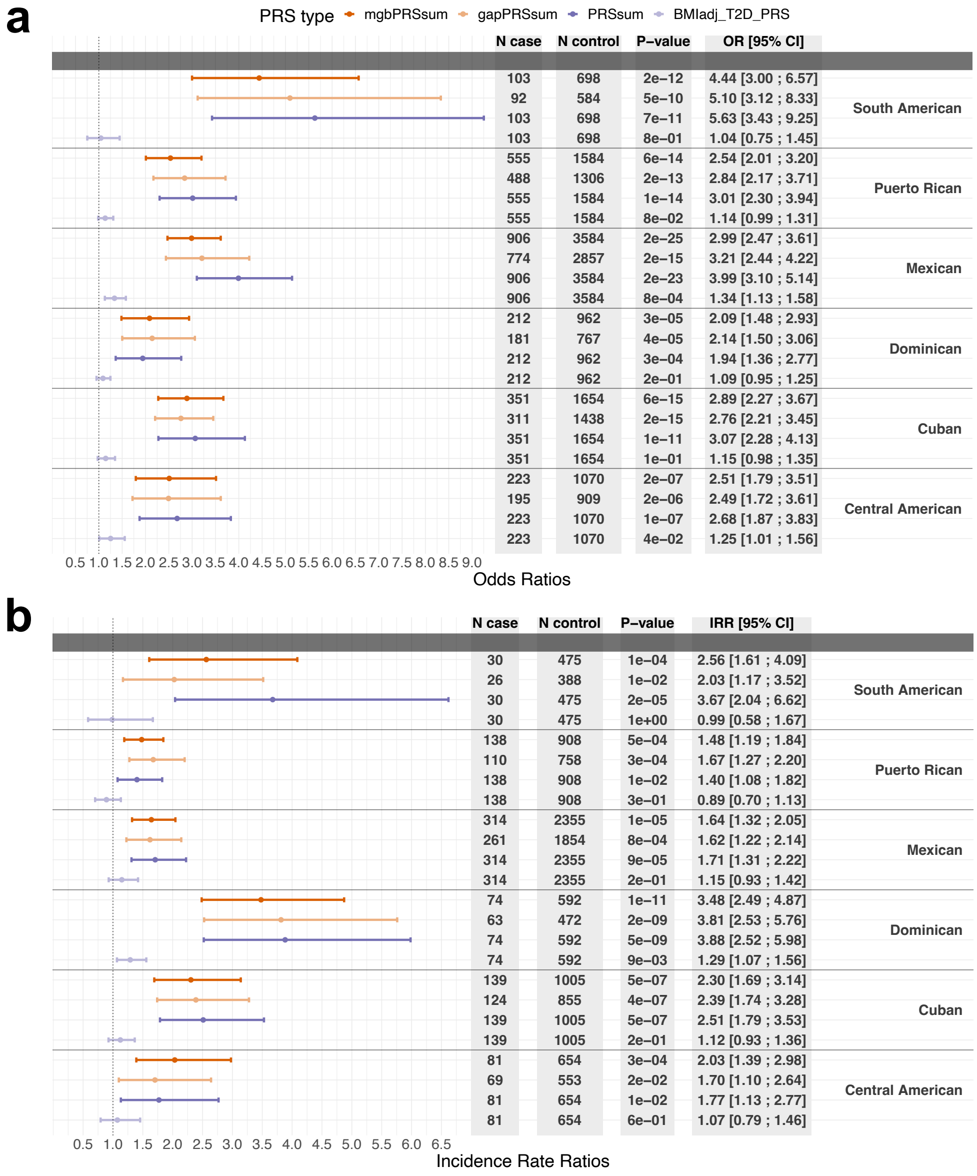


Panel a) Estimated OR of T2D-PRS in association with DM at baseline in HCHS/SOL individuals with DM status at baseline b) Estimated IRR of T2D-PRS in association with incident DM in individuals free of DM at baseline. Results are stratified by ancestral backgrounds in the overall dataset.

DM: Diabetes Mellitus; OR: odds ratio; IRR: incidence rate ratios; PRS: polygenic risk score.

Supplementary Figure 3: T2D-PRSs associations with DM stratified by OSA categories and overall dataset.

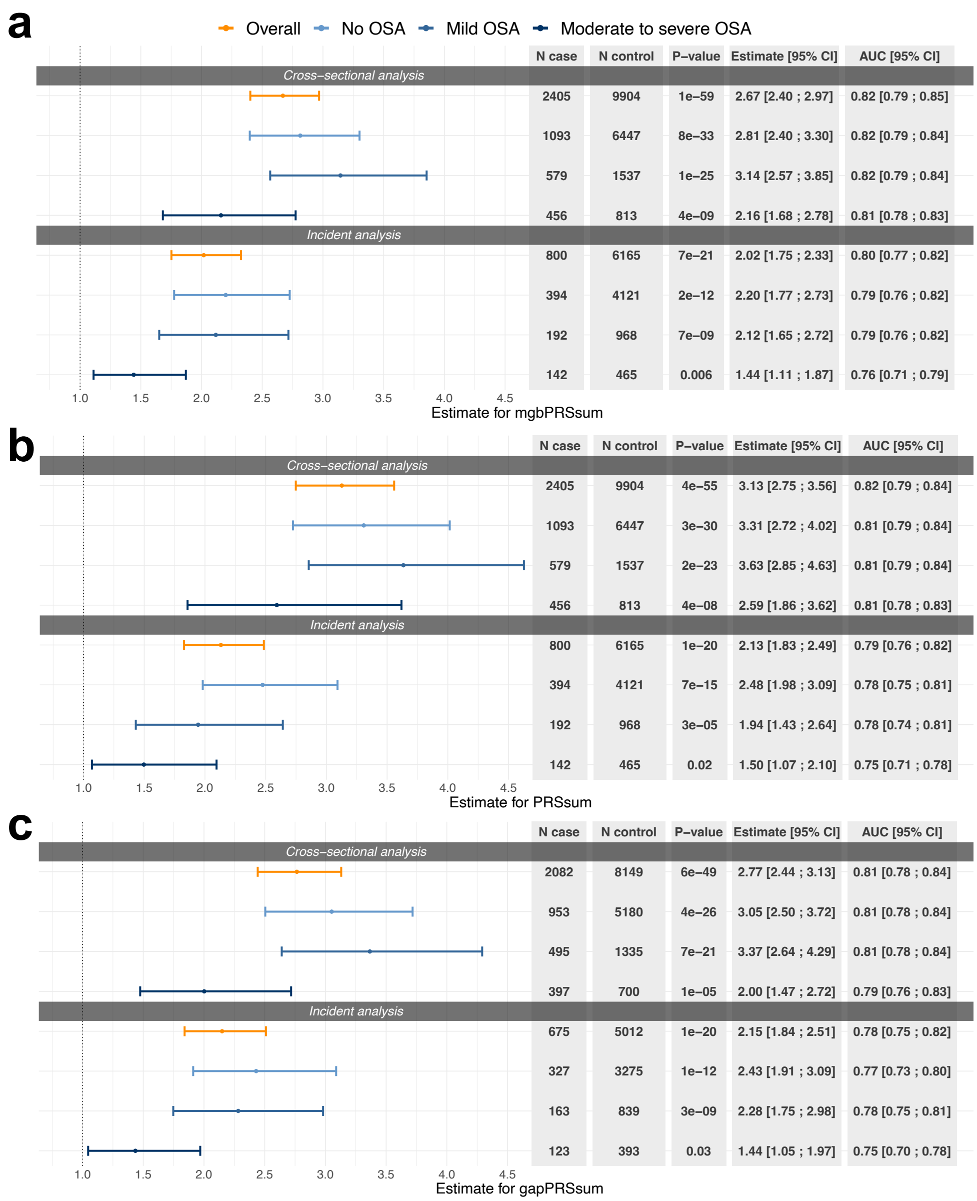


Panel **a)** Association of T2D-PRSs with DM and incident DM in HCHS/SOL individuals stratified by OSA severity levels and overall dataset. **b)** Association of PRSsum T2D-PRSs with DM and incident DM in HCHS/SOL individuals stratified by OSA categories and overall dataset. **c)** Association of gapPRSsum T2D-PRSs with DM and incident DM in HCHS/SOL individuals stratified by OSA categories and overall dataset.

All models were adjusted for age, sex, BMI, study center and 5 genetic PCs.

OR: odds ratios; IRR: incidence rate ratios; AUC: Area Under the ROC (receiver operating characteristic) Curve; T2D: type 2 diabetes; PRSs: polygenic risk scores; DM: diabetes mellitus; HCHS/SOL: Hispanic Community Health Study/Study of Latinos; EDS: excessive daytime sleepiness; OSA: obstructive sleep apnea.

#### Supplementary Figure 4: Interaction between multi-ancestry T2D-PRSs and OSA phenotypes in association with incident DM.

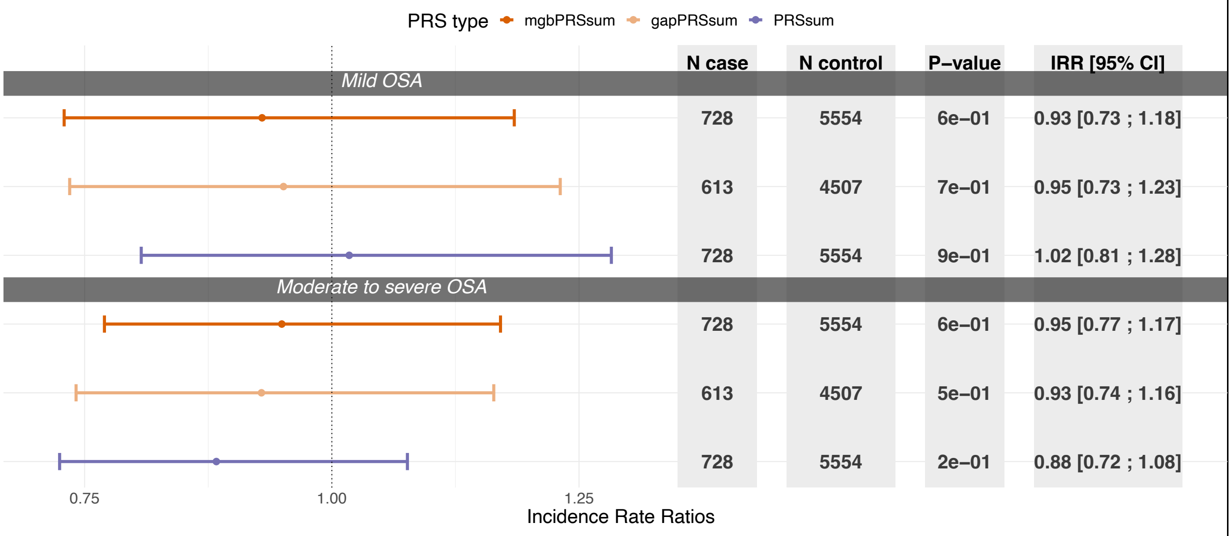


Estimated IRR of interaction between three types of T2D-PRSs and OSA in association with incident DM among normoglycemic and hyperglycemic individuals at baseline. Results are stratified by OSA severity categories.

DM: Diabetes Mellitus; IRR: incidence rate ratio; OSA: obstructive sleep apnea. PRS: polygenic risk score.

#### Supplementary Figure 5: Comparison of model performance for T2D-PRSs association with DM and incident DM using covariates and covariates plus T2D-PRS model.

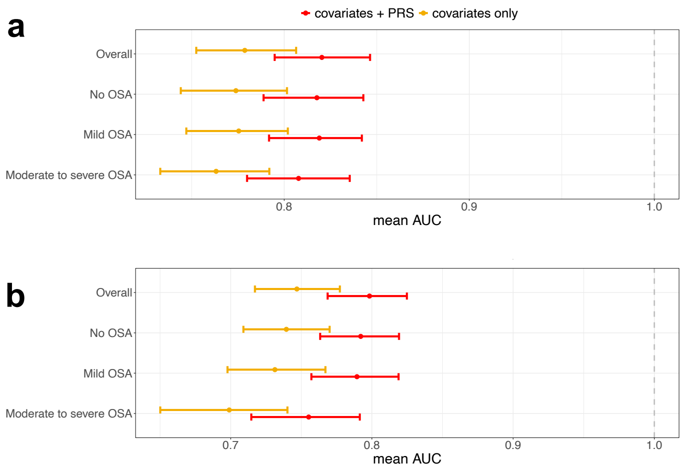


Panel **a)** Mean AUC of the "covariates model" in comparison to the performance of the "covariates plus T2D-PRS model" in estimating association between T2D-PRS and DM at baseline (testing set: overall N = 1218, no

OSA N = 1226, mild OSA N = 1254, moderate to severe OSA N = 1229). **b)** Mean AUC of the “covariates model” in comparison to the performance of the “covariates plus T2D-PRS model” in estimating association between T2D-PRS and incident DM (testing set: overall N = 877, no OSA N = 875, mild OSA N = 877, moderate to severe OSA N = 880).

AUC: area under the ROC curve; OSA: obstructive sleep apnea.

#### Supplementary Figure 6: Association of multi-ancestry T2D-PRSs with poor sleep health.

PRS type: mgbPRSsum, gapPRSsum, PRSsum, BMIadjPRS

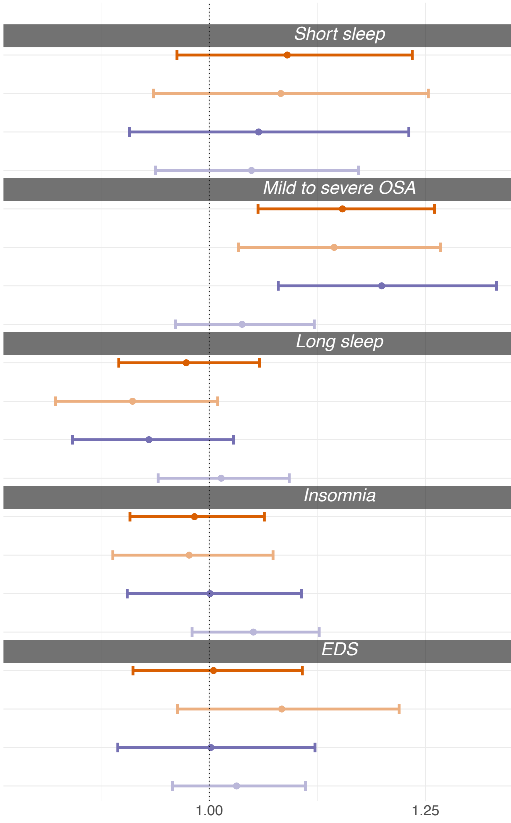


| | N case | N control | P–value | OR [95% CI] | AUC [95% CI] |
|---|---|---|---|---|---|
| *Short sleep* | | | | | |
| | 1152 | 10556 | 2e–01 | 1.09 [0.96 ; 1.23] | 0.80 [0.78 ; 0.83] |
| | 979 | 8747 | 3e–01 | 1.08 [0.94 ; 1.25] | 0.79 [0.76 ; 0.82] |
| | 1152 | 10556 | 5e–01 | 1.06 [0.91 ; 1.23] | 0.80 [0.77 ; 0.83] |
| | 1152 | 10556 | 4e–01 | 1.05 [0.94 ; 1.17] | 0.77 [0.74 ; 0.80] |
| *Mild to severe OSA* | | | | | |
| | 3344 | 7581 | 2e–03 | 1.15 [1.06 ; 1.26] | 0.82 [0.80 ; 0.84] |
| | 2888 | 6172 | 1e–02 | 1.14 [1.03 ; 1.27] | 0.81 [0.78 ; 0.84] |
| | 3344 | 7581 | 7e–04 | 1.20 [1.08 ; 1.33] | 0.82 [0.79 ; 0.84] |
| | 3344 | 7581 | 3e–01 | 1.04 [0.96 ; 1.12] | 0.78 [0.75 ; 0.81] |
| *Long sleep* | | | | | |
| | 2113 | 9595 | 5e–01 | 0.97 [0.90 ; 1.06] | 0.82 [0.79 ; 0.84] |
| | 1693 | 8033 | 8e–02 | 0.91 [0.82 ; 1.01] | 0.81 [0.78 ; 0.84] |
| | 2113 | 9595 | 2e–01 | 0.93 [0.84 ; 1.03] | 0.81 [0.79 ; 0.84] |
| | 2113 | 9595 | 7e–01 | 1.01 [0.94 ; 1.09] | 0.77 [0.74 ; 0.80] |
| *Insomnia* | | | | | |
| | 3833 | 8134 | 7e–01 | 0.98 [0.91 ; 1.06] | 0.82 [0.79 ; 0.84] |
| | 3225 | 6719 | 6e–01 | 0.98 [0.89 ; 1.07] | 0.81 [0.78 ; 0.83] |
| | 3833 | 8134 | 1e+00 | 1.00 [0.91 ; 1.11] | 0.81 [0.79 ; 0.84] |
| | 3833 | 8134 | 2e–01 | 1.05 [0.98 ; 1.13] | 0.78 [0.75 ; 0.80] |
| *EDS* | | | | | |
| | 1906 | 10206 | 9e–01 | 1.00 [0.91 ; 1.11] | 0.82 [0.79 ; 0.84] |
| | 1553 | 8510 | 2e–01 | 1.08 [0.96 ; 1.22] | 0.81 [0.78 ; 0.84] |
| | 1906 | 10206 | 1e+00 | 1.00 [0.89 ; 1.12] | 0.81 [0.78 ; 0.84] |
| | 1906 | 10206 | 4e–01 | 1.03 [0.96 ; 1.11] | 0.78 [0.75 ; 0.80] |

Estimated OR of T2D-PRS in association with poor sleep health at baseline in HCHS/SOL individuals with DM status at baseline. Results are stratified by poor sleep health categories.

DM: Diabetes Mellitus; EDS: excessive daytime sleepiness; OR: odds ratio; OSA: obstructive sleep apnea. PRS: polygenic risk score.

Supplementary Figure 7: Mediation effect of OSA on associations of mgbPRSsum with DM.

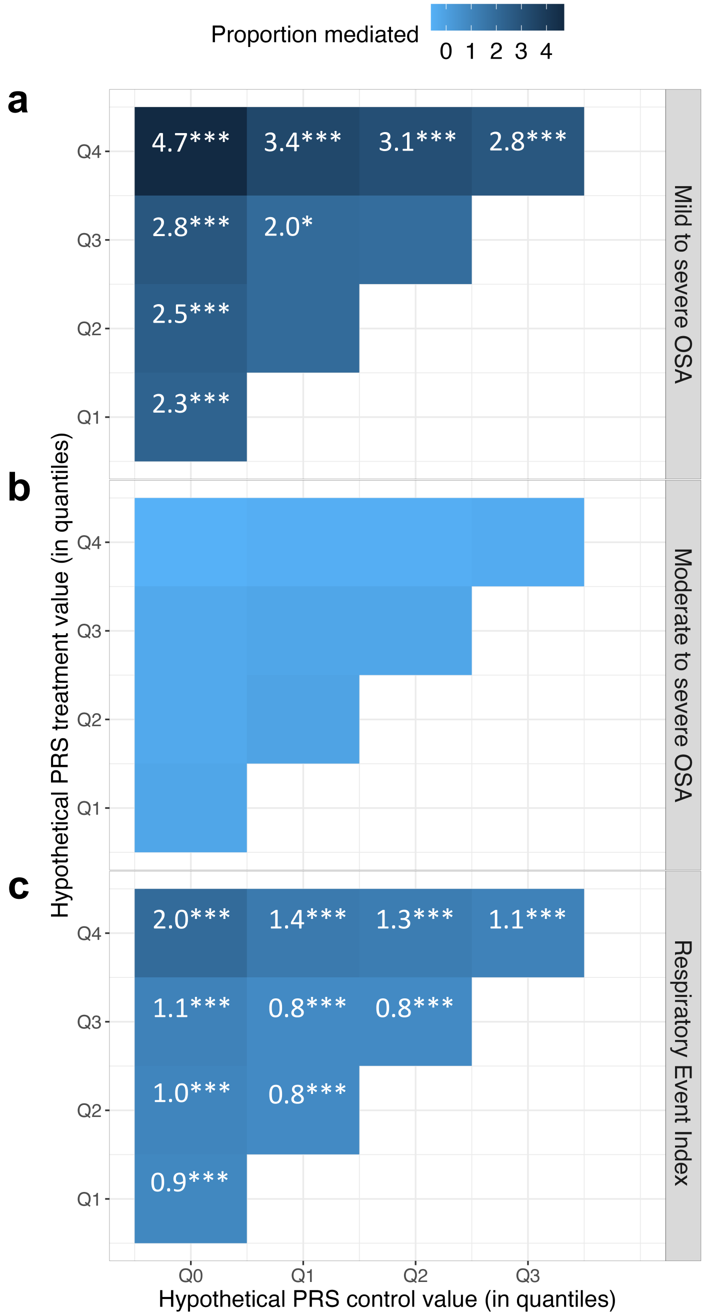


Panel **a)** Estimated proportion of mediation by the Moderate to severe OSA in the association between mgbPRSsum and incident DM in individuals who participated at the second visit to a clinic (N = 6,291). **b)** Estimated proportion of mediation by the Moderate to severe OSA in the association between mgbPRSsum and incident DM in individuals who participated at the second visit to a clinic (N = 6,291). c**)** Estimated proportion of mediation by REI in the association between mgbPRSsum and incident DM in individuals who participated at the second visit to a clinic (N = 6,291).

Significance codes: 0 >= '***' < 0.001 >= '**' < 0.01 >= '*' < 0.05 ' ' < 0.1

All models were adjusted for age, sex, BMI and 5 genetic PCs.

PRS: polygenic risk score; OSA: obstructive sleep apnea.

Supplementary Figure 8: Mediation effect of OSA on associations of BMIadjT2D-PRS with DM.

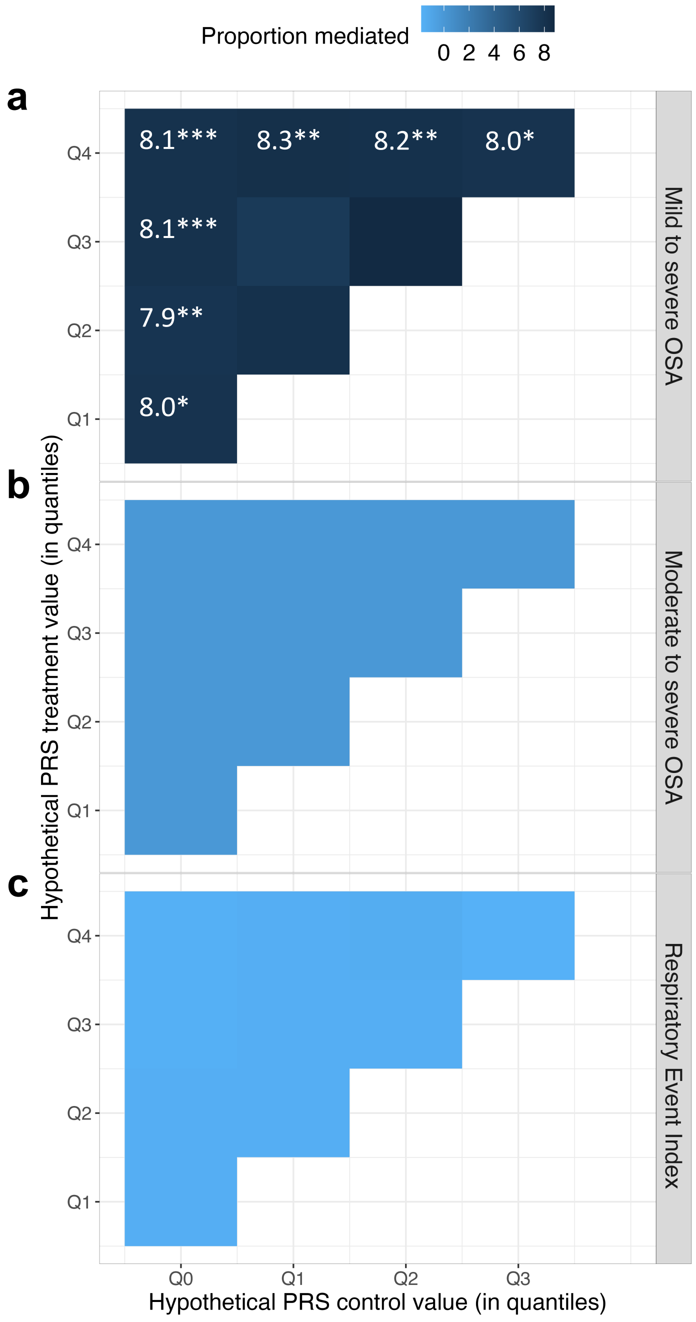


Panel **a)** Estimated proportion of mediation by the Mild to severe OSA in the association between BMIadjT2D-PRS and incident DM in individuals who participated at the second visit to a clinic (N = 6,291). **b)** Estimated proportion of mediation by the Moderate to severe OSA in the association between BMIadjT2D-PRS and incident DM in individuals who participated at the second visit to a clinic (N = 6,291). **c)** Estimated proportion of mediation by REI in the association between BMIadjT2D-PRS and incident DM in individuals who participated at the second visit to a clinic (N = 6,291).

Significance codes: 0 >= '***' < 0.001 >= '**' < 0.01 >= '*' < 0.05 ' ' < 0.1

All models were adjusted for age, sex, BMI and 5 genetic PCs.

PRS: polygenic risk score; OSA: obstructive sleep apnea.

#### Supplementary Figure 9: Estimated causal effects of OSA on T2D.

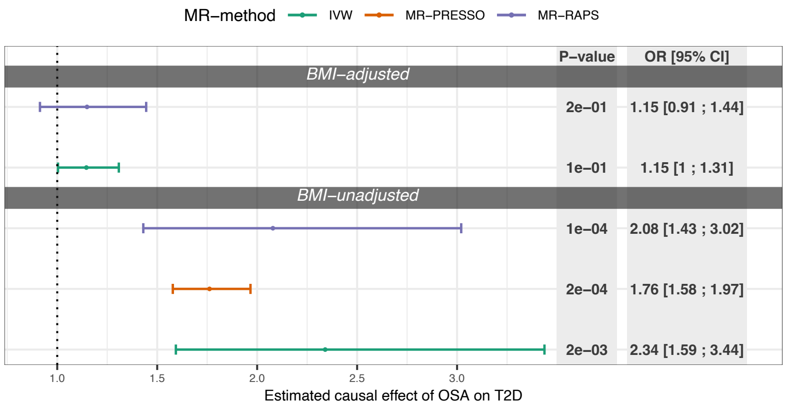


Figure: Estimated causal effect of OSA on T2D based on SNPs selected using p-value threshold < $10^{-7}$ in BMI-adjusted and BMI-unadjusted OSA GWASs.

T2D: type 2 diabetes; OSA: obstructive sleep apnea; IVW: inverse variance weighted; BMI: body mass index; OR: odds ratios;

#### Supplementary Figure 10: Association of OSA-PRS with OSA at baseline

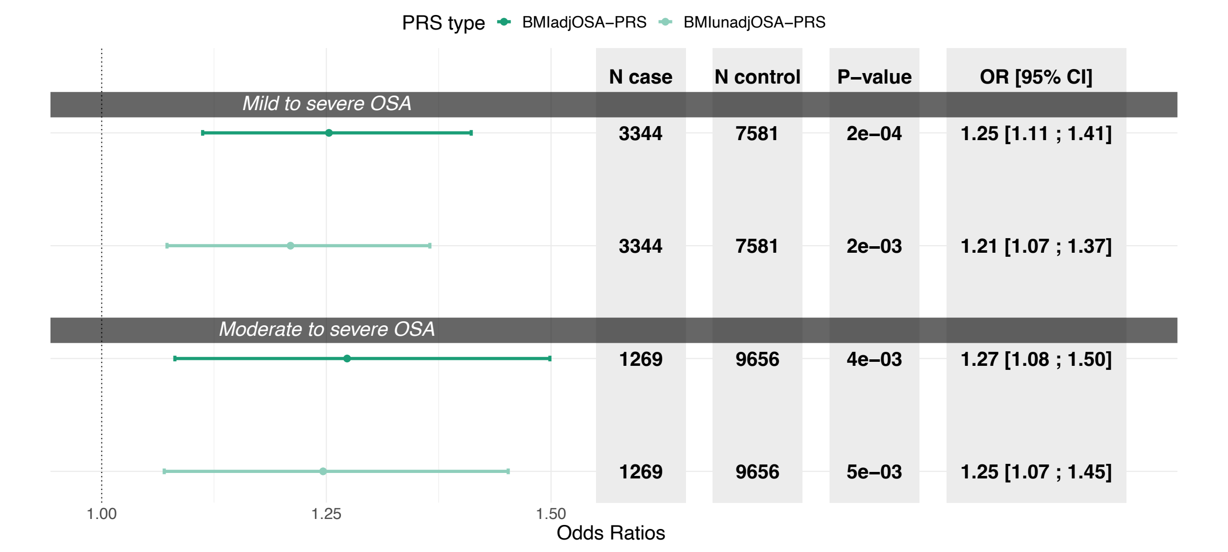


Association between OSA-PRSs (constructed using two OSA GWASs: BMI-adjusted and BMI-unadjusted) and OSA at baseline stratified by OSA severity categories: mild to severe OSA and moderate to severe.

OSA: Obstructive Sleep Apnea; PRS: polygenic risk score; BMI: body mass index; GWAS: Genome Wide Association Study; OR: Odds Ratios; CI: confidence intervals.

#### Supplementary Figure 11: Association of OSA-PRS with DM at baseline

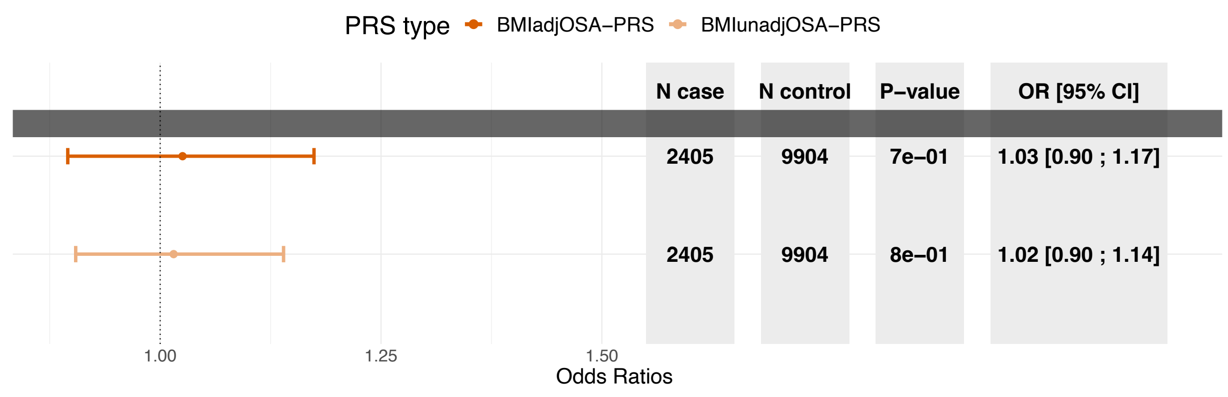


Association between OSA-PRSs (constructed using two OSA GWASs: BMI-adjusted and BMI-unadjusted) and DM at baseline in the overall dataset (N = 12,309).

OSA: Obstructive Sleep Apnea; PRS: polygenic risk score; OR: Odds Ratios; CI: confidence intervals.

#### Supplementary Note 1: the Mass General Brigham Biobank

Samples, genomic data, and health information were obtained from the Mass General Brigham (MGB) Biobank, a biorepository of consented patient samples at Mass General Brigham.

**DNA samples**

DNA samples are processed from whole blood that was collected as a dedicated research draw or as a clinical discard. Dedicated research samples are aimed to be processed within four hours of collection. Clinical discards are processed 24+ hours after collection. Whole blood is spun to buffy coat with a centrifuge and the buffy coat is stored in a freezer up to several months. The buffy coat is then extracted to DNA. The DNA is then placed in an ultralow freezer (-80$^{o}$C). Each DNA aliquot contains a minimum of 2 $\mu$g of DNA. The concentration varies.

**Genotyping**

Samples have been genotyped using three versions of the biobank SNP array offered by Illumina that is designed to capture the diversity of genetic backgrounds across the globe. The first batch of data was generated on the Multi-Ethnic Genotyping Array (MEGA) array, the first release of this SNP array. The second, third, and fourth batches were generated on the Expanded Multi-Ethnic Genotyping Array (MEGA Ex) array. All remaining data were generated on the Multi-Ethnic Global (MEG) BeadChip.

**Imputation**

Prior to performing imputation, files were converted to VCF format, separated by chromosomes. When multiple probes measured the same genotypes, they were checked for concordance and were set to a missing value if the genotypes did not match. Files were uploaded to the Michigan Imputation Server, and Genotypes were imputed using TOPMed reference panel. Genomic coordinates are provided in GRCh38.

**Quality control**

Quality control was performed using PLINK (v2.0). SNPs with low-quality imputation ($r < 0.5$), with missing call rates > 0.1, HWE p-value less than $1x10^{-6}$ and MAF <1% were filtered out. Principal components (PC) were computed using PLINK: pruning of genotype data was done using a window size of 1000 variants, sliding across the genome with a step size of 250 variants at a time, filtering out any SNPs with LD $R^2$>0.1. Computation of loadings for the first 10 PCs was done using unrelated individuals (3rd degree, identified using PLINK).

**PRS construction**

We developed polygenic scores using PRS-CSx and then PRSs were calculated with PRSice (no clumping and thresholding). All of the parameters were left at default values.

Thanks for the reminder! It looks as though scores were developed with PRS-CSx and then PRSs were calculated with PRSice. You are correct that no clumping or thresholding were performed with PRSice. All of the other parameters were left at default values.

**Diabetes status based on Curated Disease Populations**

We used the diabetes outcome from the "curated disease populations" provided by the MGB Biobank team. These phenotypes were developed by the Biobank Portal team using both structured and unstructured electronic medical record (EMR) data and clinical, computational and statistical methods. Natural Language Processing (NLP) was used to extract data from narrative text. Chart reviews by disease experts helped identify features and variables associated with particular phenotypes and were also used to validate results of the algorithms. The process produced robust phenotype algorithms that were evaluated using metrics such as sensitivity, the proportion of true positives correctly identified as such, and positive predictive value (PPV), the proportion of individuals classified as cases by the algorithm [1]. The high throughput phenotyping algorithm is as follows:

1. Create an initial phenotype definition using ICD-9 diagnosis codes.
2. Broaden the definition by determining the most up-to-date features (comorbidities, symptoms, medications) that create a more accurate profile of the phenotype when

combined with ICD-9 codes. Features are extracted from online medical literature and knowledge bases via an Automated Feature Extraction Protocol (AFEP).

3. Narrow and refine the definition by determining the features that occur most often in the Biobank data. Extract, code, and rank features contained in clinical narratives with Natural Language Processing (NLP).
4. Create a gold-standard patient set for training the method. Query coded EMR data for the set of patients having at least one ICD-9 code for the phenotype. Apply a statistical sampling algorithm to select a random subset of those patients for full chart review. A clinical expert performs a full chart review to classify the patients as positive or negative for the phenotype.
5. Train a statistical model that incorporates all features in the definition to predict the presence or absence of the phenotype against the gold-standard patient set.
6. Apply the trained model to the entire Biobank Population.

There were and 40,209 unrelated individuals. We then restricted the dataset to the individuals who are not deceased of 18 years or older reducing the data to 36,423 individuals. Of these, 2,934 (8%) had T2D according to the MAP algorithm. BMI values were taken to be the average BMI across all values available for an individual.

**Ethics statement**

All Biobank subjects have provided their consent to join the MGB Biobank, which includes agreeing to provide a blood sample linked to the electronic medical record. Subjects also agree to be recontacted by the Partners Biobank staff as needed.

**Acknowledgements**

We thank Mass General Brigham Biobank for providing samples, genomic data, and health information data.

### Supplementary Tables

#### Supplementary Table 1: GWAS summary statistics used for T2D-PRS development.

| GWAS name | Reference | Trait | Population |
|---|---|---|---|
| **DIAGRAM** | PMID: 35551307 [2] | T2D | 180,834 T2D cases and 1,159,055 controls (effective sample size 492,191) across five ancestry groups: European ancestry (51.1% of the total effective sample size); East Asian ancestry (28.4%); South Asian ancestry (8.3%); African ancestry, including recently admixed African American populations (6.6%); and Hispanic individuals with recent admixture of American, African, and European ancestry (5.6%). |
| **MVP** | PMID: 32541925 [3] | T2D | 228,499 cases and 1,178,783 controls encompassing five ancestral groups (Europeans, African Americans, Hispanics, South Asians and East Asians). MVP participants (n = 273,409) comprised predominantly male subjects (91.6%) and were classified as Europeans (72.1%), African Americans (19.5%), Hispanics (7.5%), and Asians (0.9%). |

The table provides T2D GWAS source, study population as reported by the manuscript reporting the GWAS, and number of participants used to generate summary statistics.

#### Supplementary Table 2: Characteristics of HCHS/SOL target population with no DM at baseline stratified by sleep phenotype categories.

| Characteristic | All | Healthy sleep | Insomnia | Short sleep | Long sleep | EDS | No OSA | Mild OSA | Moderate to severe OSA |
|---|---|---|---|---|---|---|---|---|---|
| **N** | **9,929** | **3,225** | **3,844** | **1,158** | **2,121** | **1,909** | **7,563** | **2,122** | **1,270** |
| **Gender N (%)** | | | | | | | | | |
| Female | 5,795 (50.5) | 1,686 (50.4) | 1,957 (59.5) | 479 (46.7) | 1,062 (54.9) | 840 (50.4) | 4,091 (54.7) | 768 (39.6) | 296 (29.2) |
| Male | 4,134 (49.5) | 1,126 (49.6) | 972 (40.5) | 410 (53.3) | 619 (45.1) | 622 (49.6) | 2,374 (45.3) | 772 (60.4) | 518 (70.8) |
| **Age** | | | | | | | | | |
| Mean (SD) | 39.23 (14.29) | 36.49 (12.96) | 42.19 (14.05) | 40.84 (14.15) | 34.97 (14.56) | 40.52 (14.66) | 36.40 (13.35) | 48.26 (12.58) | 50.21 (13.14) |
| **BMI** | | | | | | | | | |
| Mean (SD) | 28.92 (5.89) | 27.83 (5.09) | 29.59 (6.37) | 29.93 (6.02) | 28.18 (5.92) | 29.58 (6.12) | 28.04 (5.56) | 31.12 (5.81) | 33.31 (5.92) |
| **DM status at visit 1 N (%)** | | | | | | | | | |
| Normoglycemic | 5,090 (56.9) | 1,651 (57.5) | 1,387 (43.4) | 416 (42.4) | 959 (54.6) | 722 (44.3) | 3,742 (56.8) | 544 (28.8) | 216 (18.3) |
| Hyperglycemic | 4,839 (36.6) | 1,161 (33.8) | 1,542 (37.2) | 473 (41.2) | 722 (30.2) | 740 (38.2) | 2,723 (33.1) | 996 (46.8) | 598 (47.1) |
| **DM status at visit 2 N (%)** | | | | | | | | | |
| Normoglycemic | 2,437 (26.3) | 821 (30.1) | 671 (23.2) | 203 (22.6) | 434 (24.3) | 339 (25.7) | 1,845 (29.8) | 251 (17.7) | 95 (11.3) |
| Hyperglycemic | 3,735 (32.3) | 995 (30.0) | 1,173 (35.7) | 356 (35.1) | 567 (23.3) | 605 (37.7) | 2,283 (29.5) | 719 (43.0) | 369 (45.4) |
| Diabetic | 803 (6.1) | 162 (4.1) | 282 (7.4) | 68 (6.0) | 403 (13.4) | 149 (8.1) | 394 (4.4) | 192 (11.2) | 143 (14.4) |
| Missing DM status | 2,954 (35.4) | 834 (35.8) | 803 (33.7) | 262 (36.3) | 717 (39.0) | 369 (28.4) | 1,943 (36.3) | 378 (28.0) | 207 (28.8) |

#### Supplementary Table 3: Characteristics of HCHS/SOL target population at baseline stratified by sleep phenotype categories.

| Characteristic | All | Healthy sleep | Insomnia | Short sleep | Long sleep | EDS | No OSA | Mild OSA | Moderate to severe OSA |
|---|---|---|---|---|---|---|---|---|---|
| **N** | **12,342** | **3,225** | **3,844** | **1,158** | **2,121** | **1,909** | **7,563** | **2,122** | **1,270** |
| **Gender N (%)** | | | | | | | | | |
| Female | 7,244 (50.9) | 1,941 (51.0) | 2,581 (60.0) | 646 (48.4) | 1,340 (55.1) | 1,102 (50.3) | 4,816 (55.4) | 1,117 (42.4) | 508 (31.9) |
| Male | 5,098 (49.1) | 1,284 (49.0) | 1,263 (40.0) | 512 (51.6) | 781 (44.9) | 807 (49.7) | 2,747 (44.6) | 1,005 (57.6) | 762 (68.1) |
| **Age** | | | | | | | | | |
| Mean (SD) | 41.51 (15.05) | 37.58 (13.49) | 44.70 (14.53) | 43.46 (14.80) | 37.94 (15.98) | 43.03 (15.07) | 37.87 (14.03) | 50.46 (12.83) | 52.62 (12.91) |
| **BMI** | | | | | | | | | |
| Mean (SD) | 29.40 (6.13) | 28.06 (5.19) | 30.21 (6.70) | 30.49 (6.28) | 28.75 (6.35) | 30.28 (6.70) | 28.30 (5.66) | 31.47 (6.05) | 33.71 (6.25) |
| **DM status at visit 1 N (%)** | | | | | | | | | |
| Normoglycemic | 5,090 (48.4) | 1,651 (57.5) | 1,387 (43.4) | 416 (42.4) | 959 (54.6) | 722 (44.3) | 3,742 (56.8) | 544 (28.8) | 216 (18.3) |
| Hyperglycemic | 4,839 (36.6) | 1,161 (33.8) | 1,542 (37.2) | 473 (41.2) | 722 (30.2) | 740 (38.2) | 2,723 (33.1) | 996 (46.8) | 598 (47.1) |
| Diabetic | 2,413 (15.0) | 413 (8.8) | 915 (19.4) | 269 (16.4) | 440 (15.2) | 447 (17.4) | 1,098 (10.1) | 582 (24.4) | 456 (34.6) |
| **DM status at visit 2 N (%)** | | | | | | | | | |
| Normoglycemic | 2,462 (22.5) | 826 (27.5) | 679 (18.9) | 203 (18.8) | 434 (24.3) | 344 (21.6) | 1,854 (26.9) | 258 (13.6) | 98 (7.5) |
| Hyperglycemic | 3,891 (28.6) | 1,023 (28.1) | 1,232 (30.4) | 368 (30.2) | 567 (23.3) | 631 (32.2) | 2,364 (27.4) | 757 (34.8) | 392 (31.5) |
| Diabetic | 2,438 (14.7) | 451 (9.6) | 891 (17.7) | 254 (16.2) | 403 (13.4) | 462 (18.1) | 1,160 (10.5) | 584 (24.0) | 445 (31.0) |
| Missing DM status | 3,551 (34.2) | 925 (34.8) | 1,042 (33.0) | 333 (34.8) | 717 (39.0) | 472 (28.1) | 2,185 (35.2) | 523 (27.6) | 335 (30.0) |

#### Supplementary Table 4: Characteristics of MGB dataset stratified by T2D status.

| Characteristic | No T2D | T2D | Overall |
|---|---|---|---|
| **N** | **33,489** | **2,934** | **36,423** |
| **Gender N (%)** | | | |
| Female | 19,022 (56.8%) | 1,325 (45.2%) | 20,347 (55.9%) |
| Male | 14,467 (43.2%) | 1,609 (54.8%) | 16,076 (44.1%) |
| **Age** | | | |
| Mean (SD) | 57.7 (17.0) | 68.6 (12.6) | 58.6 (17.0) |
| Median [Min, Max] | 60.0 [21.0, 104] | 70.0 [25.0, 101] | 61.0 [21.0, 104] |
| **Self-reported background** | | | |
| White | 28,252 (84.4%) | 2,120 (72.3%) | 30,372 (83.4%) |
| Black | 1,473 (4.4%) | 354 (12.1%) | 1,827 (5.0%) |
| Hispanic | 1,099 (3.3%) | 202 (6.9%) | 1,301 (3.6%) |
| Asian | 692 (2.1%) | 61 (2.1%) | 753 (2.1%) |
| American Indian/Alaskan Native | 42 (0.1%) | 8 (0.3%) | 50 (0.1%) |
| More than one | 309 (0.9%) | 21 (0.7%) | 330 (0.9%) |
| Unknown | 1,622 (4.8%) | 168 (5.7%) | 1,790 (4.9%) |
| **BMI** | | | |
| Mean (SD) | 28.2 (8.01) | 32.8 (7.32) | 28.6 (8.05) |
| Median [Min, Max] | 27.1 [2.66, 748] | 31.9 [11.7, 173] | 27.5 [2.66, 748] |
| Missing | 3,614 (10.8%) | 182 (6.2%) | 3,796 (10.4%) |

#### Supplementary Table 5: Characteristics of HCHS/SOL target population stratified by self-reported Hispanic background.

| Characteristic | Central American | Cuban | Dominican | Mexican | Puerto Rican | South American | More than one/Other heritage | Overall |
|---|---|---|---|---|---|---|---|---|
| **N** | **1,295** | **2,009** | **1,177** | **4,502** | **2,149** | **803** | **382** | **12,317** |
| **Gender N (%)** | | | | | | | | |
| Female | 767 (59.2%) | 1,053 (52.4%) | 772 (65.6%) | 2,715 (60.3%) | 1,246 (58.0%) | 471 (58.7%) | 203 (53.1%) | 7,227 (58.7%) |
| Male | 528 (40.8%) | 956 (47.6%) | 405 (34.4%) | 1,787 (39.7%) | 903 (42.0%) | 332 (41.3%) | 179 (46.9%) | 5,090 (41.3%) |
| **Age** | | | | | | | | |
| Mean (SD) | 44.8 (13.4) | 49.3 (13.0) | 45.6 (14.3) | 44.6 (13.8) | 48.2 (14.0) | 47.0 (13.2) | 39.6 (15.2) | 46.1 (13.9) |
| **BMI** | | | | | | | | |
| Mean (SD) | 29.9 (5.86) | 29.2 (5.82) | 29.4 (5.76) | 29.8 (5.91) | 30.9 (6.89) | 28.7 (5.21) | 30.0 (6.46) | 29.8 (6.06) |
| Missing N (%) | 2 (0.2%) | 4 (0.2%) | 3 (0.3%) | 12 (0.3%) | 10 (0.5%) | 2 (0.2%) | 0 (0%) | 33 (0.3%) |
| **DM status at baseline N (%)** | | | | | | | | |
| Diabetic | 223 (17.2%) | 352 (17.5%) | 212 (18.0%) | 910 (20.2%) | 557 (25.9%) | 104 (13.0%) | 48 (12.6%) | 2,406 (19.5%) |
| Hyperglycemic | 516 (39.8%) | 846 (42.1%) | 438 (37.2%) | 1,770 (39.3%) | 802 (37.3%) | 323 (40.2%) | 133 (34.8%) | 4,828 (39.2%) |
| Normoglycemic | 556 (42.9%) | 811 (40.4%) | 527 (44.8%) | 1,822 (40.5%) | 790 (36.8%) | 376 (46.8%) | 201 (52.6%) | 5,083 (41.3%) |
| **Incident DM N (%)** | | | | | | | | |
| No incident | 252 (19.5%) | 398 (19.8%) | 255 (21.7%) | 903 (20.1%) | 378 (17.6%) | 182 (22.7%) | 91 (23.8%) | 2,459 (20.0%) |
| Incident | 646 (49.9%) | 1,010 (50.3%) | 575 (48.9%) | 2,481 (55.1%) | 1,062 (49.4%) | 410 (51.1%) | 137 (35.9%) | 6,321 (51.3%) |
| Missing | 397 (30.7%) | 601 (29.9%) | 347 (29.5%) | 1,118 (24.8%) | 709 (33.0%) | 211 (26.3%) | 154 (40.3%) | 3,537 (28.7%) |

#### Supplementary References


1. Yu, S., et al., *Toward high-throughput phenotyping: unbiased automated feature extraction and selection from knowledge sources.* J Am Med Inform Assoc, 2015. **22**(5): p. 993-1000.
2. Mahajan, A., et al., *Multi-ancestry genetic study of type 2 diabetes highlights the power of diverse populations for discovery and translation.* Nat Genet, 2022. **54**(5): p. 560-572.
3. Vujkovic, M., et al., *Discovery of 318 new risk loci for type 2 diabetes and related vascular outcomes among 1.4 million participants in a multi-ancestry meta-analysis.* Nat Genet, 2020. **52**(7): p. 680-691.
